# Antidepressant Response in Major Depressive Disorder: A Genome-wide Association Study

**DOI:** 10.1101/2020.12.11.20245035

**Authors:** Oliver Pain, Karen Hodgson, Vassily Trubetskoy, Stephan Ripke, Victoria S. Marshe, Mark J. Adams, Enda M. Byrne, Adrian I. Campos, Tania Carrillo-Roa, Annamaria Cattaneo, Thomas Damm Als, Daniel Souery, Mojca Z. Dernovsek, Chiara Fabbri, Caroline Hayward, Neven Henigsberg, Joanna Hauser, James L. Kennedy, Eric J. Lenze, Glyn Lewis, Daniel J. Müller, Nicholas G. Martin, Benoit H. Mulsant, Ole Mors, Nader Perroud, David J. Porteous, Miguel E. Rentería, Charles F. Reynolds, Marcella Rietschel, Rudolf Uher, Eleanor M. Wigmore, Wolfgang Maier, Naomi R. Wray, Katherine J. Aitchison, Volker Arolt, Bernhard T. Baune, Joanna M. Biernacka, Guido Bondolfi, Katharina Domschke, Masaki Kato, Qingqin S. Li, Yu-Li Liu, Alessandro Serretti, Shih-Jen Tsai, Gustavo Turecki, Richard Weinshilboum, the GSRD Consortium, the Major Depressive Disorder Working Group of the Psychiatric Genomics Consortium, Andrew M. McIntosh, Cathryn M. Lewis

## Abstract

**Importance:** Antidepressants are a first line treatment for depression. However, only a third of individuals remit after the first treatment. Genetic variation likely regulates antidepressant response, yet the success of previous genome-wide association studies has been limited by sample size.

**Objective:** Gain insight into underlying biology of antidepressant response, characterize SNP-based heritability and genetic overlap with related outcomes, and evaluate out-of-sample prediction using polygenic scores.

**Design:** Genome-wide meta-analysis of antidepressant response measures, Remission and Percentage Improvement in depression scores.

**Setting:** Multiple international recruitment sites, including clinical trial and open label studies.

**Participants:** Diagnosed with Major Depressive Disorder and assessed for depressive symptoms before and after prescription of an antidepressant medication.

**Main Outcome(s) and Measure(s):** Antidepressant response measured as Remission and Percentage Improvement.

**Results:** Genome-wide analysis of Remission (*N_remit_*=1,852, *N_non-remit_*=3,299) and Percentage Improvement (*N*=5,218) identified no genome-wide significant variants. The heritability from common variants was significantly different from zero for Remission (*h*^2^=0.132, SE=0.056), but not Percentage Improvement (*h*^2^=-0.018, SE=0.032). Polygenic score analysis showed better antidepressant response was associated with lower genetic risk for schizophrenia, and higher genetic propensity for educational attainment. Polygenic scores for antidepressant response demonstrated weak but statistically significant evidence of out-of-sample prediction across cohorts, though results varied in external cohorts.

**Conclusions and Relevance:** This study demonstrates antidepressant response is influenced by common genetic variation, has a genetic overlap with schizophrenia and educational attainment, and provides a useful resource for future research. Larger sample sizes are required to attain the potential of genetics for understanding and predicting antidepressant response.

**Key Points:** *Question:* What is the genetic architecture of antidepressant response, and how is it associated with other traits?

*Findings:* This genome-wide association study of antidepressant response finds Remission SNP-based heritability was significantly different from zero for Remission (*h*^2^=0.132, SE=0.056), but not Percentage Improvement (*h*^2^=-0.018, SE=0.032). Polygenic score analysis showed better antidepressant response was associated with lower genetic risk for schizophrenia, and higher genetic propensity for educational attainment.

*Meaning:* This study demonstrates antidepressant response is influenced by common genetic variation, has a genetic overlap with schizophrenia and educational attainment, and provides a useful resource for future research.

## Introduction

Major depressive disorder (MDD) is the third leading cause of years lived with disability world-wide(1) and a substantial risk factor for suicide(2). MDD confers a major personal, societal and economic burden(3), partly because of the limited efficacy of treatment options.

In 2011–2014, 12.7% of individuals in the United States aged 12 and over reported antidepressant medication use(4). The rate of antidepressant prescriptions is also increasing, with the number of prescriptions doubling in the United Kingdom in the decade prior to 2018(5). Antidepressants are robustly linked to a reduction in depressive symptoms(6) but they are often ineffective: approximately 35% of patients remit after their primary treatment(7) and approximately 40% develop treatment resistant depression, defined as not remitting after two or more antidepressants(8). For patients, the process of trialing antidepressants can be lengthy and demoralizing, delaying recovery and exposing patients to a range of potential side-effects that reduce adherence and willingness to try new drugs(9). There is therefore great potential to improve treatment of depression through better understanding of the factors that control response to antidepressants and implementing this knowledge through individually tailored treatment.

Pharmacogenetic studies were expected to uncover loci with large effects on drug response and adverse events due to effects of pharmacokinetic or pharmacodynamic mechanisms. Whilst associations between antidepressant plasma levels and drug metabolizing enzymes CYP2D6 and CYP2C19 have been identified(10–12), previous research suggests that genes encoding these enzymes and other candidate genes account for a small proportion of variation in drug response(13,14). However, genotyping complexities for such candidate genes may contribute to limited findings.

Several genome-wide association studies (GWAS) have been performed to identify genetic predictors of antidepressant response. Although no robustly replicated associations have been detected to date(15–19), common single nucleotide polymorphisms (SNPs) are reported to explain 42% (SE=18%, 95%CI=7%-77%) of the variance(20). Pharmacogenetic studies are intensive to perform, requiring disease severity measures at baseline pre-treatment and then longitudinally, with many studies being performed as part of a randomized controlled trial(15–18). This ‘clinically assessed’ approach provides high quality data, though it has led to previous studies being limited in sample size, with less than 3,000 MDD patients in the largest GWAS to date. Further efforts to combine these individual cohorts to increase sample size for genetic studies are therefore required. Use of lighter phenotyping approaches such as electronic health record derived treatment resistant depression (TRD)(21) may also provide novel insight, though it is unclear whether these different measures of antidepressant response have a common genetic basis.

In this study, we analyse genome-wide genetic data on clinically assessed antidepressant response from 5,843 patients treated for MDD, combined from 13 international research studies. Using this novel data resource, we perform GWAS of Remission and Percentage Improvement after receiving antidepressant medication, and undertake extensive post-GWAS analyses, made feasible through this increased sample size. This study aims to elucidate the genetic architecture of antidepressant response and use polygenic scores to establish the relationship between antidepressant response and mental health outcomes. We find, for the first time, a replicable polygenic signal of antidepressant response across studies.

## Methods

### Primary Samples and Measures

This study analysed 13 cohorts (Table 1). Ten cohorts were of European ancestry, and three were of East Asian ancestry (Supplementary Material). All subjects provided written informed consent for pharmacogenetic analyses. These primary cohorts include individuals with a clinical diagnosis of MDD, assessed for depressive symptoms before and after treatment with antidepressants.

**Table 1.** Cohorts of individuals diagnosed with MDD and assessed for depressive symptoms at before and after treatment with antidepressant medication.

| Study | Country, Region | Study design | Study length (wks) | Medications | Measure | Age (Median) | Age (IQR) | Percentage females | N | N perc. improv. | N remit | N non-remit |
| --- | --- | --- | --- | --- | --- | --- | --- | --- | --- | --- | --- | --- |
| **European ancestry** |  |  |  |  |  |  |  |  |  |  |  |  |
| **STARD** (45) | USA | Open label | 12 | Citalopram | QIDSC | 44 | 21 | 58% | 1163 | 1163 | 506 | 657 |
| **GSRD** (17) | Europe | Naturalistic | >4 | Various | MADRS | 52.5 | 18 | 66% | 1152 | 1152 | 189 | 963 |
| **GENDEP** (46) | Europe | Partially randomized RCT | 12 | Escitalopram, Nortriptyline | MADRS | 43 | 18 | 63% | 783 | 783 | 291 | 365 |
| **DAST** | Germany | Naturalistic inpatient | 6 | Various | HAMD-21 | 50 | 25 | 57% | 586 | 586 | 245 | 303 |
| **PGRN-AMPS** (47) | USA | Open label | 8 | Citalopram, Escitalopram | QIDSC | 38.5 | 21 | 63% | 490 | 392 | 200 | 290 |
| **GENPOD** (18) | UK | Open label | 12 | Citalopram, Reboxetine | BDI | 38 | 18 | 69% | 474 | 474 | 169 | 305 |
| **PFZ** (18) | USA | RCT | 6-8 | Sertraline, Fluoxetine, Paroxetine | HAMD-17 | 43 | 22 | 67% | 309 | 309 | 99 | 210 |
| **MAYO** (16) | USA | Open label | 8 | Citalopram, Escitalopram | HAMD-17 | 37 | 22 | 62% | 156 | 156 | 80 | 76 |
| **GSK** (18) | USA | RCT | 8 | Escitalopram | HAMD-17 | 36 | 19.25 | 55% | 132 | 132 | 56 | 76 |
| **GODS** (18) | Switzerland | Open label | 8 | Paroxetine | MADRS | 37 | 14 | 52% | 71 | 71 | 17 | 54 |
| **East Asian ancestry** |  |  |  |  |  |  |  |  |  |  |  |  |
| **MIAOLI**<br>(16) | Taiwan | Open label | 8 | Escitalopram,<br>Paroxetine | HAMD-17 | 41 | 22 | 82% | 233 | 233 | 103 | 130 |
| **TAIPEI**<br>(16) | Taiwan | Open label | 8 | Fluoxetine,<br>Citalopram | HAMD-17 | 46 | 25 | 55% | 174 | 174 | 45 | 129 |
| **JAPAN**<br>(16) | Japan | RCT | 6 | Fluvoxamine,<br>Paroxetine | HAMD-17 | 44.5 | 24 | 47% | 120 | 120 | 78 | 42 |
| **Total** |  |  |  |  |  |  |  |  | **5843** | **5745** | **2078** | **3600** |

RCT: randomized control trial
Age (IQR): Interquartile range of age
*N*: Number of participants included after quality control of genetic and clinical data
*N* perc. improv.: Number of participants with percentage improvement information
*N* remit: Number of participants that remitted
*N* non-remit: Number of participants that did not remit
QIDSC: Quick Inventory of Depressive Symptomatology
BDI: Beck Depression Inventory
MADRS: Montgomery Asberg Depression Rating Scale
HAMD-17: Hamilton Depression Rating Scale 17 item scale

Two measures of antidepressant response were defined, Remission and Percentage Improvement. Remission is a binary measure attained when a patient’s depression symptom score decreases to a pre-specified threshold for the rating scale (Supplementary material).

All analyses included covariates of the first 20 principal components of population structure, age and gender. Analyses using the Remission measure of response also included baseline symptom score as a covariate, to control for depression severity.

Each cohort underwent standard quality control and 1000 Genomes Phase 3 imputation using the RICOPILI pipeline on the LISA server(22) (Supplementary material and Table S1).

### Genome-wide Association Study (GWAS)

GWAS was performed using the RICOPILI pipeline(22) separately for studies with participants of European and of East Asian ancestry (Supplementary Material). All other analyses were performed using only the European ancestry cohorts due to the limited sample size of the East Asian cohorts. See Supplementary Material for description of gene-level and gene-set enrichment analysis.

### Estimation of SNP-based heritability

The SNP-based heritability of Remission and Percentage Improvement was estimated using individual-level data by GREML (genomic-relatedness-based restricted maximum-likelihood) in the software GCTA (Genome-wide Complex Trait Analysis)(23,24). The analysis was performed firstly, across all cohorts, including a study covariate (mega-GREML), and secondly, separately within each cohort and then inverse variance meta-analyzed (meta-GREML) (Supplementary Material). Comparison of mega- and meta-GREML estimates can provide insight into the heterogeneity between cohorts, since only mega-GREML accounts for genetic covariances between cohorts.

### Leave-one-out polygenic scoring

To determine whether polygenic scores derived from the Remission and Percentage Improvement GWAS summary statistics predict antidepressant response in an independent sample, a leave-one-out polygenic scoring approach was used. This involves calculating polygenic scores within each cohort based on GWAS summary statistics derived using all other cohorts. Polygenic scores were calculated using PRSice V2(25) (Supplementary Material). One-sided *p*-values were used to assess statistical significance as we are testing the one-sided hypothesis that the polygenic score has a positive association with the outcome in the target sample.

### Estimation of genetic overlap with mental health phenotypes

We tested for evidence of genetic overlap between antidepressant response measures and seven mental health phenotypes: major depression(26), bipolar disorder(27), schizophrenia(28), attention deficit hyperactivity disorder (ADHD)(29), autism spectrum disorder (ASD)(30), anxiety(31), and problematic drinking (Alcohol Use Disorders Identification Test (AUDIT) problem subscale)(32). Educational attainment(33) was also included as it has strong correlations with the mental health disorders tested. Evidence of genetic overlap was assessed using polygenic scoring with AVENGEME(34), and LDSC(35). Several cohorts (STAR*D, GENPOD, GENDEP, PFZ) of the major depression susceptibility GWAS were included in this antidepressant response study and AVENGEME results should be interpreted with caution.

AVENGEME aggregates polygenic scores association results across p-value threshold to estimate genetic covariance between antidepressant response and the eight mental-health phenotypes. AVENGEME parameters are provided in Table S2. Bonferroni correction was used to account for multiple testing for the eight discovery GWAS used.

### Replication cohorts and analyses

#### Out-of-sample prediction

External validation of polygenic scores derived using the full GWAS results was also carried out. Five independent samples were used (Supplementary Material). In brief, Janssen (N=190, remission rate=11.8%)(36), Douglas Biomarker Study (N=127, remission rate=23.6%)(37), and IRL-GREY (N=307, remission rate=52.4 %)(38) prospectively assessed depressive symptoms, concordant with the discovery GWAS samples. In contrast, Generation Scotland (*N*_treatment-resistent_=177, *N*_Non-treatment-resistant_=2,455)(21) assessed electronic prescription data, and the Australian Genetics of Depression Study (AGDS; *N*_responders_=4,368, *N*_Non-responders_=6,879)(39) collected retrospective self-report questionnaire data. Polygenic score association results were meta-analyzed across the prospectively assessed cohorts given their more comparable study design and antidepressant measures. One-sided *p*-values were used to assess statistical significance.

#### Comparison of genetic covariance with mental health phenotypes

Individual-level data were available for Generation Scotland enabling estimation of genetic covariance between TRD and mental health-related phenotypes using AVENGEME, as described above. Analyses in Generation Scotland were controlled for age, gender, and 20 principal components of population structure. Individuals with depression in Generation Scotland were also included in the major depression GWAS so results should be interpreted with caution.

## Results

Descriptive statistics for the cohorts used in this study are available in Table 1 and Figures S1-S5.

### Genome-wide association study of antidepressant response

Across the 10 European studies, 5,151 individuals with Remission data (1,852 patients remitting (36.0%)) and 5,218 participants with Percentage Improvement data were available. No variants were significantly associated with Remission or Percentage Improvement (Figures S6-S7, Tables S4-S5). There was no evidence of confounding (Figures S8-S9, Table S6)

No significant associations were identified in the East Asian GWASs (N=527, Figures S10-S11). A comparison between East Asian and European GWAS results shown in Supplementary material. See Supplementary Material for gene-level and gene-set enrichment analysis results.

### SNP-based heritability

Analysis across all samples (mega-GREML) showed Remission to have a significant non-zero SNP-based heritability (*h*^2^=0.132; SE=0.056; 95%CI=0.022-0.241; *p*=0.009, liability scale assuming population prevalence of 0.3), whereas the SNP-based heritability for Percentage improvement was not significantly different from zero (*h*^2^=-0.018; SE=0.032; 95%CI=-0.080-0.045; *p*=0.303)(Figure 1).

**Figure 1.**
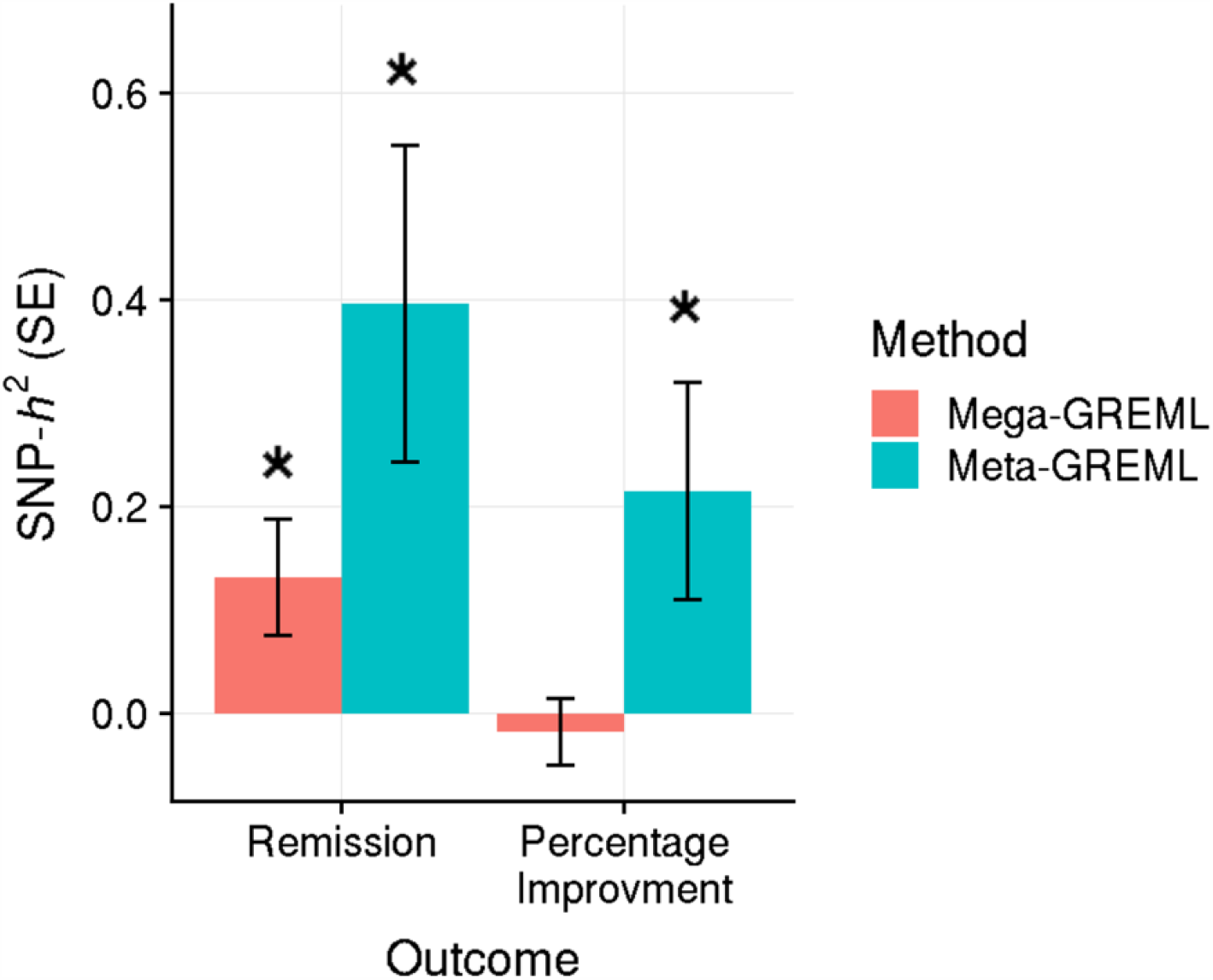
SNP-based heritability (SNP-*h*^*2*^) estimates for Remission and Percentage Improvement with standard error bars. Figure shows across (mega-) and within (meta-) sample GREML estimates. * *indicates estimate is significantly different from zero, at* p<0.05.

The SNP-based heritability estimates from meta-analysis of within-sample estimates (meta-GREML) were significant for both Remission (*h*^2^=0.396; SE=0.153; 95%CI=0.096-0.696; *p*=0.010, liability scale assuming population prevalence of 0.3) and Percentage Improvement (*h*^2^=0.215; SE=0.105; 95%CI=0.009-0.421; *p*=0.041) (Figure 1). See Supplementary Material for SNP-based heritability sensitivity analyses.

### Out-of-sample prediction

Leave-one-out polygenic score analysis provided evidence that polygenic scores derived using Remission and Percentage Improvement GWAS results could both explain a statistically significant amount of variance out-of-sample (Figure 2). Both Remission and Percentage Improvement explained ∼0.1% of the variance, with polygenic scores for multiple *p*-value thresholds associated at nominal significance.

**Figure 2.**
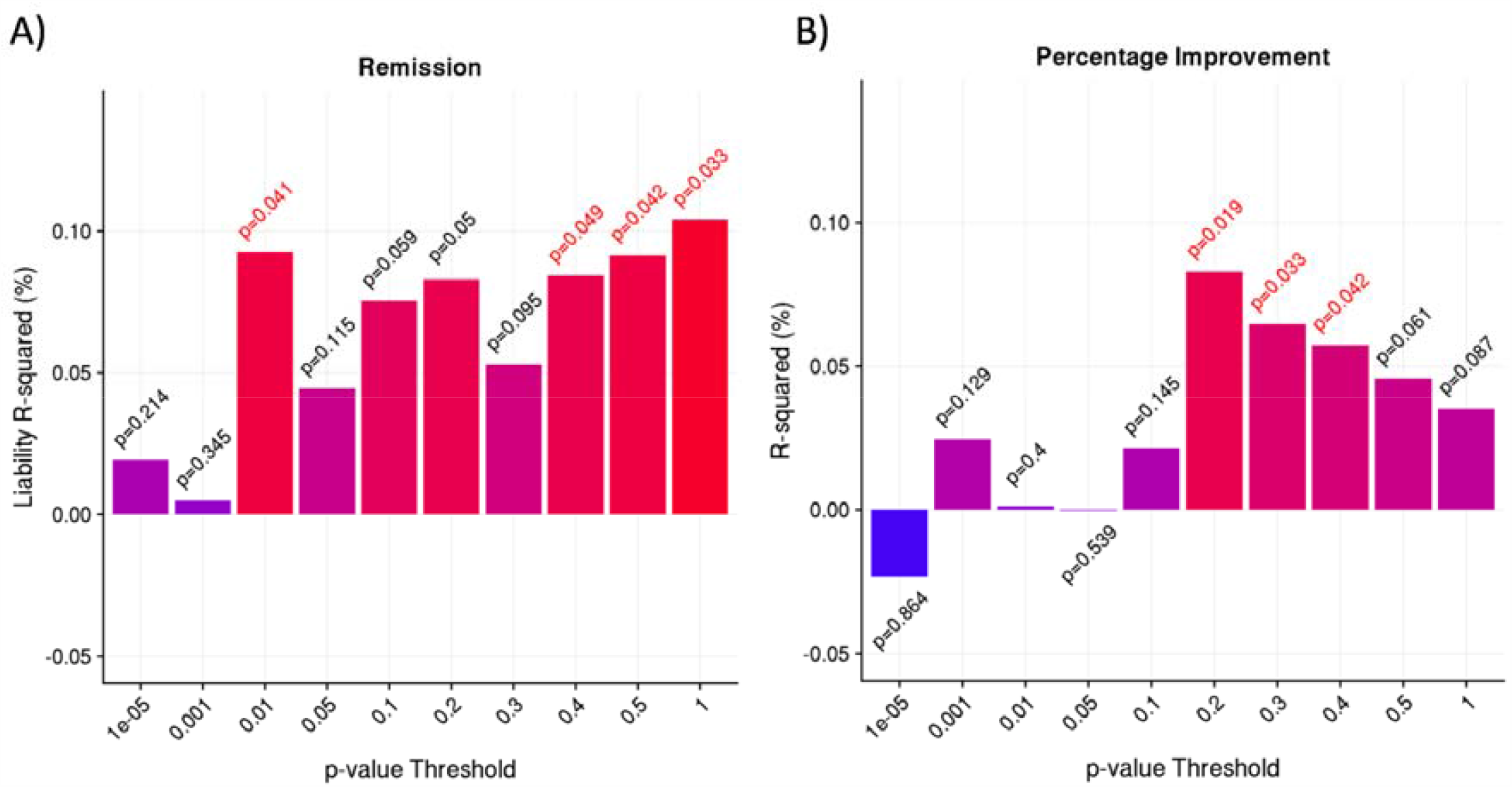
Polygenic prediction of antidepressant response from leave-one-out polygenic scoring for A) Remission and B) Percentage Improvement. R^2^ estimates are signed to indicate positive or negative association. One-sided p-values are shown above or below the bars, with p-values < 0.05 highlighted in red.

Validation of polygenic scores based on the full antidepressant response GWAS summary statistics was carried out using five samples. Meta-analysis of polygenic score associations across the three prospectively-assessed cohorts (Janssen, Douglas Biomarker Study and IRL-GREY) showed nominally significant evidence of association for the Remission polygenic score (maximum liability *r*^2^=0.8%, *p*-value=0.015), and a non-significant association for the Percentage Improvement score (maximum *r*^2^=0.2%, *p*-value=0.091) (Figure S19). Results were highly variable across each prospectively-assessed cohort. No association was found between polygenic scores in Generation Scotland or AGDS cohorts. Full polygenic score replication results are in Tables S10-S13.

### Genetic overlap with mental health phenotypes

Both Remission and Percentage Improvement showed a significant negative genetic covariance with schizophrenia, and significant positive genetic covariance with educational attainment (Figure 3, Tables S14-S15). Percentage Improvement also showed a significant negative covariance with bipolar disorder and a significant positive genetic covariance with ASD. LDSC genetic correlation estimates were broadly concordant, although non-significant (Figure S20).

**Figure 3.**
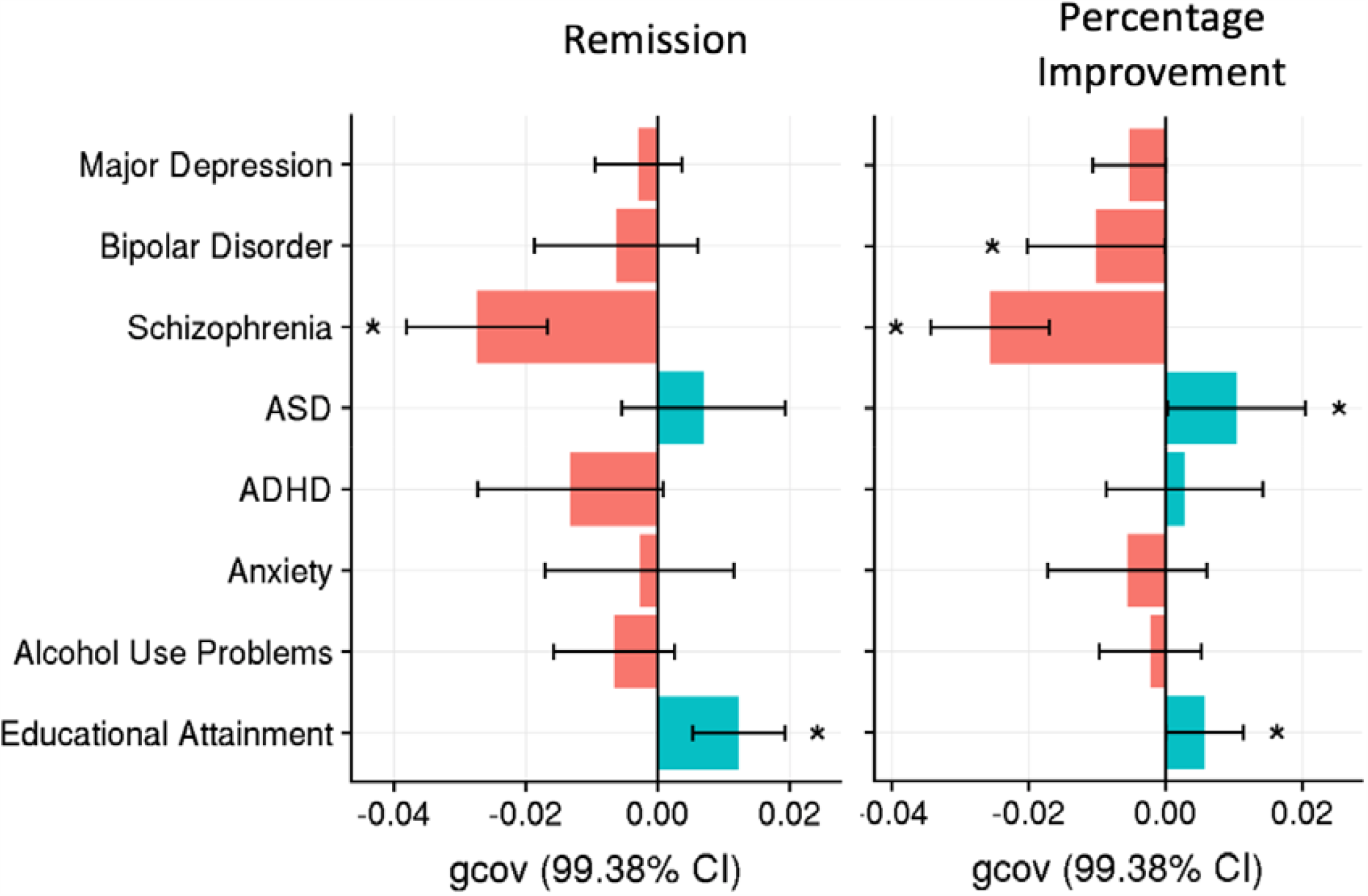
Genetic covariance estimates between antidepressant response phenotypes and seven mental health phenotypes and educational attainment. gcov = genetic covariance; 99.38% CI = Confidence intervals corrected for multiple testing.

Genetic overlap estimates between TRD in Generation Scotland and mental health phenotypes were congruent with results from primary samples, showing genetic risk for schizophrenia was greater among individuals with TRD, and educational attainment genetic propensity was greater among individuals that were non-TRD (Figure S21).

## Discussion

Antidepressants are a common and effective strategy for treating MDD; however, remission rates are typically low and factors affecting antidepressant response are poorly understood. This study is the largest genetic investigation of antidepressant response based on clinically defined cohorts. For the first time, we identify a polygenic profile for antidepressant response, which can predict across cohorts, and shows genetic correlations with traits that reflect clinical observations.

This study finds significant evidence that antidepressant response is influenced by common genetic variation. Meta-analysis of SNP-based heritability estimates within each cohort indicate that 20-40% of the variance in antidepressant response is attributable to common genetic variation, consistent with a previous analysis of a subset of these studies(20). However, the SNP-based heritability decreased substantially when estimating across cohorts simultaneously. These results indicate antidepressant response in a broad context has a heritable component, but genetic differences can explain additional variability in antidepressant response within more specific contexts. Despite the apparent heterogeneity across individual cohorts, the sample sizes for antidepressant response are sufficiently large to detect a polygenic signal. Genetic studies for susceptibility to psychiatric disorders show that findings accrue after an inflection point in sample size is reached(26–28). This study’s findings for SNP-based heritability and out-of-sample polygenic prediction indicate that sample sizes for antidepressant response are reaching the inflection point and that larger studies will uncover more of the genetic component(40). Power calculations for detecting genome-wide significant variation, and the variance explained by corresponding polygenic scores, are provided in Supplementary Figure 22. Interestingly, our findings suggest the SNP-based heritability of Remission is higher than for Percentage Improvement. This needs to be investigated further but it may be that Percentage Improvement is more susceptible to random variation in depressive symptoms or because it can also capture increases in depressive symptoms over time.

This study provides novel insight into the shared genetic basis between antidepressant response and mental health phenotypes. We show an association between high genetic liability of psychiatric disorders and poorer response, which mirrors conclusions of clinical studies(41). The schizophrenia polygenic risk score was negatively associated with antidepressant response, which is replicated in the TRD phenotype in Generation Scotland. Previous studies have shown that individuals with TRD may respond to antipsychotic medication(42). Our findings extend those reports by suggesting that individuals with antidepressant resistance also have a higher burden of schizophrenia genetic risk. In contrast, we report a novel finding that high ASD genetic liability increased the chance of remission.

Another recent study reported ASD genetic liability is associated with poorer response to cognitive behavioral therapy (CBT)(43). If both these findings are replicated, it would suggest ASD genetic liability could serve as a differential predictor of response to antidepressants and CBT. We also identified a significant association between genetic propensity for educational attainment and improved antidepressant response as well as non-TRD. This may reflect the indirect measurement of socioeconomic status captured by educational attainment, which is supported by previous literature showing a positive association between antidepressant response and socioeconomic status(44). Future research should explore whether individuals with higher educational attainment have improved response due to factors such as adherence or joint psychological treatment.

Polygenic scores derived from the Remission and Percentage Improvement GWASs both significantly predicted antidepressant response out-of-sample using a leave-one-out design. This is the first GWAS of antidepressant response able to predict significantly out-of-sample, representing an important advance in the field of antidepressant response genetics. Although the variance explained is low (*r*^2^ = 0.1%) and *p*-values are close to the nominal significance threshold, this result is encouraging given the sample size of this study. For example, a recent GWAS of MDD explains only 1.9% of the variance in MDD, despite having a sample size 100 times greater than this study(26). Our finding suggests that a renewed effort to systematically collect new samples in which genetic associations with antidepressant response can be identified will improve the prediction of antidepressant response, helping to uncover its biological mechanisms and clinical associations, and eventually enable more accurate clinical predictors to be developed and applied.

In addition, no association was detected with genetic variation within classical pharmacokinetic candidate genes, such as *CYP2D6* and *CYP2C19*, which have previously been robustly associated with antidepressant plasma levels(11). Although the enzymatic activity of CYP2D6 and CYP2C19 is largely regulated by common genetic variation, these variants include structural variants that are not well captured by GWAS arrays, and large effects on enzymatic activity are typically conferred by combinations of genetic variants (haplotypes), which GWAS does not assess. Therefore, the absence of an association at this point may be a false negative result. Furthermore, looking across individuals that have not been treated with a specific antidepressant or antidepressant class will reduce the likelihood of detecting pharmacokinetic effects.

Due to a limited sample size it was not possible to estimate genetic correlations between longitudinally assessed antidepressant response and TRD defined using electronic health records. However, comparison of shared genetic etiology with other mental health phenotypes indicated these distinct measures of antidepressant response have a shared genetic basis. Further comparison and integration of these two approaches is warranted and may prove fruitful given the large gains in sample size that electronic health record derived phenotypes can provide.

There are several limitations to this study that should be addressed in the future. First, large sample sizes are essential for robust identification of associated genetic variation and out-of-sample prediction. However, combining independently collected datasets, inevitably introduces heterogeneity. Obtaining large homogenous samples is particularly challenging for pharmacogenetic studies as heterogeneity is not only driven by patient characteristics such as diagnosis and patient ascertainment, but also by differences in treatment such as the drug, dosage, duration and co-pharmacotherapy. Although the cohorts within this study have many features in common, heterogeneity in antidepressant treatment is present. As sample sizes grow, analyses stratified by these factors will become more feasible, enabling detection of genetic effects specific to antidepressants and other treatment characteristics. Second, an important question to consider is whether the variance in depressive symptoms after treatment is due to antidepressant response or other variables altering the course of depression. Although antidepressants have a significant effect on depressive symptoms and their administration is the core feature of participants in this study, individuals may vary in depressive symptoms due to other factors affecting disease progression, such as clinical and socio-demographic variables and placebo response. This is a difficult issue to resolve but should be considered when interpreting the results. Future genetic studies incorporating the placebo arm of clinical trials may help identify genetic associations specific to antidepressant response. Third, this study has focused on changes in total depressive symptoms without considering symptom domain-specific changes or the presence of side effects. Given the wide range of depressive symptoms and the influence side effects can have on efficacy, consideration of these features may provide additional insights. Fourth, although this study included three cohorts of East Asian ancestry, further inclusion of cohorts with diverse ancestries is an important area. Genetic analysis within diverse populations helps to ensure the findings are applicable to worldwide populations and can help fine-map causal variants underlying genetic associations.

In summary, this study identifies a polygenic profile for antidepressant response that predicts across studies and is negatively correlated with genetic susceptibility to schizophrenia, which could be used for prognostic purposes. Whilst the current results have no clinical utility as a pharmacogenetic test, they indicate that studies with larger sample sizes could provide predictions explaining a substantial proportion of antidepressant response. We note that a prognostic test that enables even a modest increase in the proportion of patients that respond to antidepressants would have a substantial impact on recovery for many patients, given the high prevalence of depression. We hope this study prompts both replication and extension to accelerate the development of pharmacogenetic testing for psychiatry.

## Data Availability

Access to summary statistics will be made available through the Psychiatric Genomics Consortium on publication of the peer-reviewed manuscrtip.

https://www.med.unc.edu/pgc/

## Disclosures

Cathryn M. Lewis is on the scientific advisory board for Myriad Neuroscience. Katherine J. Aitchison has received two research grants in the last two years from Janssen Inc., Canada (fellowship grants for trainees) and provided consultancy services in the last three years for Otsuka Canada Pharmaceutical Inc., Lundbeck Canada, and HLS Therapeutics. Alessandro Serretti is or has been consultant/speaker for: Abbott, Abbvie, Angelini, Astra Zeneca, Clinical Data, Boheringer, Bristol Myers Squibb, Eli Lilly, GlaxoSmithKline, Innovapharma, Italfarmaco, Janssen, Lundbeck, Naurex, Pfizer, Polifarma, Sanofi, Servier. Andrew M. McIntosh has received research support from the Sackler Trust and speakers fees from Janssen and Illumina. Masaki Kato has received grant funding from Japan Society for the Promotion of Science, SENSHIN Medical Research Foundation and Japan Research Foundation for Clinical Pharmacology, and speaker’s honoraria from Dainippon-Sumitomo Pharma, Otsuka, Meiji-Seika Pharma, Eli Lilly, MSD K.K., GlaxoSmithkline, Pfizer, Janssen Pharmaceutical, Shionogi, Mitsubishi Tanabe Pharma, Takeda Pharmaceutical, Lundbeck and Ono Pharmaceutical. Qingqin S. Li is an employee of Janssen Research & Development, LLC and a shareholder in Johnson & Johnson, the parent company of the Janssen companies. Qingqin S. Li declares that, except for income received from her primary employer, no financial support or compensation has been received from any individual or corporate entity over the past three years for research or professional service, and there is no personal financial holding that could be perceived as constituting a potential conflict of interest.

## Acknowledgements

*GSRD:* The collection of this sample was supported by an unrestricted grant from Lundbeck for the Group for the Study of Resistant Depression (GSRD). Lundbeck had no further role in the study design, in the collection, analysis and interpretation of data.

*GENDEP:* GENDEP was funded by the European Commission Framework 6 grant, EC Contract Ref.: LSHB-CT-2003-503428. H. Lundbeck provided nortriptyline and escitalopram for the GENDEP study. GlaxoSmithKline and the UK National Institute for Health Research of the Department of Health contributed to the funding of the sample collection at the Institute of Psychiatry, London. GENDEP Illumina array genotyping was funded in part by a joint grant from the U.K. Medical research council and GlaxoSmithKline (G0701420).

*GENPOD:* GENPOD was funded by the Medical Research Council and supported by the Mental Health Research Network, UK. The genotyping of GENPOD samples was supported by the Innovative Medicine Initiative Joint Undertaking (IMI-JU) under grant agreement n° 115008 of which resources are composed of European Union and the European Federation of Pharmaceutical Industries and Associations (EFPIA) in-kind contribution and financial contribution from the European Union’s Seventh Framework Programme (FP7/2007-2013). EFPIA members Pfizer, Glaxo Smith Kline, and F. Hoffmann La-Roche have contributed work and samples to the project presented here.

*PFZ, GSK and GODS*: Supported by the Innovative Medicine Initiative Joint Undertaking (IMI-JU) under grant agreement n° 115008 of which resources are composed of European Union and the European Federation of Pharmaceutical Industries and Associations (EFPIA) in-kind contribution and financial contribution from the European Union’s Seventh Framework Programme (FP7/2007-2013). EFPIA members Pfizer, Glaxo Smith Kline, and F. Hoffmann La-Roche have contributed work and samples to the project presented here.

*PGRN-AMPS:* PGRN-AMPS data was obtained via dbGAP (accession number: phs000670.v1.p1). Funding support for the Pharmacogenomics Research Network Antidepressant Medication Pharmacogenomic Study (PGRN-AMPS) was provided by the National Institute of General Medical Sciences (NIGMS), National Institutes of Health (NIH), through the PGRN grant to Principal Investigators R. Weinshilboum and L. Wang (U19 GM61388). Dr. D. Mrazek served as the Principal Investigator for the PGRN-AMPS study within the Mayo Clinic PGRN program. Genome-wide genotyping was performed at the RIKEN Center for Genomic Medicine, with funding provided by RIKEN. The datasets used for the analyses described in this manuscript were obtained from dbGaP at http://www.ncbi.nlm.nih.gov/projects/gap/.

*Generation Scotland:* Generation Scotland is supported by the Wellcome Trust (reference 104036/Z/14/Z, 216767/Z/19/Z), The Chief Scientist Office of the Scottish Government Health Department (CZD/16/6) and the Scottish Funding Council (HR03006). Data linkage and analysis in Generation Scotland is supported by the Medical Research Council (MC_PC_17209).

*Douglas Biomarker Study:* The genotyping of the samples was funded and generated by Janssen Research & Development, LLC

*PGC:* We are deeply indebted to the investigators who comprise the PGC, and to the hundreds of thousands of subjects who have shared their life experiences with PGC investigators. Major funding for the PGC is from the US National Institutes of Health (U01 MH109528 and U01 MH109532). Statistical analyses were carried out on the NL Genetic Cluster Computer (http://www.geneticcluster.org) hosted by SURFsara.

*KCL:* Authors OP and CML are funded by the National Institute for Health Research (NIHR) Biomedical Research Centre at South London and Maudsley NHS Foundation Trust and King’s College London. The authors acknowledge support from the Medical Research Council (MR/N015746/1) and use of the research computing facility at King’s College London, Rosalind (https://rosalind.kcl.ac.uk), which is delivered in partnership with the NIHR Maudsley BRC, and part-funded by capital equipment grants from the Maudsley Charity (award 980) and Guy’s & St. Thomas’ Charity (TR130505). The views expressed are those of the authors and not necessarily those of the NHS, the NIHR or the Department of Health and Social Care.

## Notes

### Author Declarations

All studies recieved ethical approval from the relevant ethics committee or institutional review board. The ethical details of each study are given in the Supplementary Materials in the Ethical Information table, p3-5.

