## Supplemental Materials for "Antidepressant Response in Major Depressive Disorder: A Genome-wide Association Study"

**Supplementary Material for Investigating the Common Genetic Basis of Antidepressant Response**

**Table of figures:**

### Supplementary material

**Ethical information**

Summary ethical information for each cohort included in the meta-analysis is listed below. All studies received approval from the relevant institutional review board. Ethical permission for this meta-analysis study was obtained from King’s College London Research Ethics Office PNM REP (LRS-18/19-11645).

| **Cohort or sample** | **Abbreviation** | **Pubmed ID** | **Ethics** |
| --- | --- | --- | --- |
| STAR*D | stard | 19846067 | The University of Texas Southwestern Medical Center at Dallas and the institutional review boards at each clinical site and regional center and the Data Coordinating Center and the Data Safety and Monitoring Board of the National Institute of Mental Health (NIMH) approved and monitored the protocol. Written consent was obtained from all participants after the procedures and any associated risks were explained. ClinicalTrials.gov identifier (NCT number): NCT00021528 |
| GSRD | gsrd | 30468137 | All procedures involved in the study were approved by the local ethics committees of each participating centre (coordinating centre approval number: B406201213479). Further details can be found elsewhere ^9^. Written informed consent was obtained from all patients included in this study. |
| GENDEP | gendep | 20360315 | The study was approved by ethics boards in all participating centres. The main GENDEP study received Ethical Approval from the Institute of Psychiatry Research Committee on 2nd June 2004, Approval Number 292/03. All participants provided a written consent after the procedures were explained. GENDEP is registered at EudraCT (No.2004-001723-38, http://eudract.emea.europa.eu) and ISRCTN (No. 03693000, http://www.controlledtrials.com). |
| DAST | dast | NA | Approval of the ethics committee of the University of Muenster, Muenster, Germany, and written informed consent from all subjects were obtained before commencement of any study procedures. |
| PGRN-AMPS | pgrn | 22907730 | All patients provided written informed consent. The study protocol was approved by the Mayo Clinic Institutional Review Board. |
| GENPOD | genpod | 23091423 | Ethical approval was obtained from the South West Ethics Committee (MREC 02/6/076) as well as research governance approval from Bristol, Manchester and Newcastle Primary Care NHS Trusts. The ISRCTN is 31345163 and EudraCT number 2004-001434-16. |
| Pfizer | pfz | 23091423 | Analysis was based on participants from eight MDD clinical trials (n=345). All study protocols received institutional review board (IRB) approval and informed consent was obtained from participating subjects prior to sample collection. |
| MAYO | mayo | 25897834 | The Mayo cohort was collected as a continuation of the PGRN-AMPS cohort (above). All patients provided written informed consent. The study protocol was approved by the Mayo Clinic Institutional Review Board. |
| GSK | gsk | 23091423 | Institutional review boards^22^ approved the protocols for each of the study sites for these randomized double-blind, double-dummy, placebo-controlled, multicentre studies (study 1 Feb 2003 – June 2004: GSK protocol AK130926; study 2 Jan 2003 – June 2004; GSK protocol AK130926). |
| GODS | gods | 23091423 | The study protocol was approved by the ethics committee of the Geneva University Department of Psychiatry and written informed consent was obtained from all subjects. |
| MIAOLI | mioali | 25897834 | This study was approved by the institutional review boards of the National Health Research Institutes, Taiwan and all participating clinics, with the requirement of written informed consent signed by each study participant. The study was assigned ClinicalTrials.gov Identifier NCT00384020. |
| TAIPEI | taipei | 25897834 | The study was approved by the Institutional Review Board of Taipei Veterans General Hospital (VGHIRB No.: 95-11-07) and E-DA hospital (E-MRP-095-014), and written informed consent was obtained from the participants prior to enrolment. |
| JAPAN | japan | 25897834 | The study was approved by the ethical committee of the Kansai Medical University and Osaka University. Written informed consent was obtained from each subject after a detailed explanation of the study. |

#### Primary samples information

**STAR*D**

The STAR*D study is a trial of protocol-guided antidepressant treatment for outpatients with major depression ^1^. The study included 4,041 treatment-seeking adult outpatients, recruited in 18 primary care and 23 psychiatric clinical sites across the United States. Inclusion criteria were a diagnosis of non-psychotic unipolar major depressive disorder diagnosed by a clinician and confirmed with a checklist of DSM-IV criteria; age 18 to 75 years; and a minimum score of 14 on the HAMD (Hamilton Depression Rating Scale). The present meta-analysis uses data from the first treatment step, which included protocol-guided citalopram (20–60 mg/day) ^2^. Depression severity in STAR*D was rated every 2 weeks using the clinician-rated and self-report versions of the 16-item Quick Inventory for Depressive Symptomatology (QIDS) ^3^. The primary outcome measure was the 17-item HAMD (HAMD-17), administered by trained independent evaluators at study entry and at the end of each treatment step ^1^. However, since data from QIDS were available for more participants and this assessment tool was found to be closely equivalent to HAMD, most STAR*D reports rely on it primarily ^2,4^. Genetic material was collected from 1,948 (48%) participants; of whom 1,491 (37% of the original STAR*D sample, including 980 of white/European ancestry) passed quality control and were included in previously reported genome-wide analyses ^5^. The study was approved by institutional ethics review boards at all centres. Written consent was obtained from all participants after the procedures and any associated risks were explained. STAR*D genotype and phenotype data are available through the National Institute of Mental Health Human Genetic Initiative ([https://www.nimhgenetics.org/](about:blank)).

**GSRD**

The European group for the study of resistant depression (GSRD) participants were recruited within a multicentre, cross-sectional study including 1,346 adults who were in- and out-patients with MDD according to DSM-IV-TR criteria ^6^. The GSRD has been active for more than 20 years in the field of clinical and genetic modulators of TRD. Diagnosis was confirmed using the Mini International Neuropsychiatric Interview (MINI) ^7^. Inclusion criteria were: (a) having received at least one antidepressant during the current MDD episode (≥4 weeks at adequate dose); (b) Montgomery–Åsberg Depression Rating Scale (MADRS) ^8^ score >22 at the onset of the current MDD episode. Exclusion criteria were (a) any other primary psychiatric disorder than MDD, (b) any substance disorder (except nicotine and caffeine) in the previous 6 months, and (c) any condition that could interfere with the ability to give informed consent or with the assessments required by the study (for example linguistic barrier). Depressive symptom severity was assessed using the MADRS at study inclusion and at the onset of the current MDD episode. Information on previous and current antidepressant and other pharmacological treatments during the current MDD episode was collected as well as clinical–demographic characteristics. Antidepressant treatment was naturalistic according to best-clinical practice principles. All procedures contributing to this work comply with the ethical standards of the relevant national and institutional committees on human experimentation and with the Helsinki Declaration of 1975, as revised in 2008. All procedures involved in the study were approved by the local ethics committees of each participating centre (coordinating centre approval number: B406201213479). Further details can be found elsewhere ^9^. Written informed consent was obtained from all patients included in this study.

**GENDEP**

GENDEP was a twelve-week single-blind part-randomized multi-centre study with two active pharmacological treatment arms ^10^. This includes 57 additionally recruited participants which were not included in initial reports. This enlarged GENDEP sample includes 868 treatment-seeking adults (men n=321; women n=547) diagnosed with ICD-10/DSM-IV unipolar major depression of at least moderate severity established in the Schedules for Clinical Assessment in Neuropsychiatry (SCAN) interview ^11^. Severity of depression was assessed weekly by three established rating scales ^12^. Personal or family history of bipolar disorder or schizophrenia and active substance dependence constituted exclusion criteria. Individuals were of white European origin and ranged from 19 to 72 years of age with a mean age of 42.6 years (S.D=11.7). Eligible participants were treated with either escitalopram or nortriptyline. Patients with no contraindications were randomly allocated to flexible-dosage nortriptyline (50–150 mg daily) or escitalopram (10–30 mg daily) for 12 weeks. Patients with contraindications for one drug were offered the other. A total of 628 participants (77%) completed at least 8 weeks of treatment with the originally allocated antidepressant ^10^. Individuals were excluded from the analysis if they had no post baseline information. The study was approved by ethics boards in all participating centres. All participants provided a written consent after the procedures were explained. GENDEP is registered at EudraCT (No.2004-001723-38, http://eudract.emea.europa.eu) and ISRCTN (No. 03693000, http://www.controlledtrials.com).

**DAST**

Parts of the present sample have already been described in published genetic or pharmacogenetic studies targeting other gene systems ^13–15^. Briefly, patients were ascertained from consecutive admissions for inpatient treatment at the Department of Psychiatry, University of Muenster, Germany, between 2004 and 2008. Patients’ lifetime diagnoses related to major depression and bipolar disorder were ascertained by the use of a structured clinical interview (SCID-I) according to the criteria of DSM-IV ^6^. Patients with schizoaffective disorders or comorbid substance abuse disorders, mental retardation, pregnancy and neurological, neurodegenerative disorders or other clinically unstable medical illnesses impairing psychiatric evaluation were not included in this analysis. Clinical course of depression was assessed using the 21-item HAMD (HAMD-21) scale on a weekly basis. Patients were treated in a naturalistic setting with a variety of antidepressant medication according to doctor’s choice. None of the included patients had received electroconvulsive therapy within six months before the present investigation. Approval of the ethics committee of the University of Muenster, Muenster, Germany, and written informed consent from all subjects were obtained before commencement of any study procedures.

**PGRN-AMPS**

The Mayo Clinic Pharmacogenomic Research Network Antidepressant Medication Pharmacogenomic Study (PGRN-AMPS) was supported by the NIGMS-Pharmacogenomics Research Network (PGRN), which has been described elsewhere ^16,17^. The PGRN-AMPS is an ongoing eight week outpatient SSRI clinical trial that was performed at the Mayo Clinic in Rochester, MN. Patients enrolled in the study met diagnostic criteria for MDD without psychosis or mania and had a HAMD-17 score ≥14. The study was designed with inclusion and exclusion criteria similar to those used in the Sequenced Treatment Alternatives to Relieve Depression study (STAR*D) ^18^. Potential study subjects taking an antidepressant, antipsychotic or mood stabilizing medication were not eligible for enrollment. Patients with MDD initially received either 10 mg of escitalopram or 20 mg of citalopram. SSRI efficacy was determined using the 16-item Quick Inventory of Depressive Symptomatology (QIDS-C16) and HAMD-17 scores after four weeks and then eight weeks of SSRI therapy. At four weeks after the initiation of treatment, the dose could be increased to 20 mg of escitalopram or 40 mg of citalopram after a clinical assessment of the subject. Unless there was a contraindication, dose was increased if the QIDS-C16 score at the follow-up visit was ≥9, and possibly following a clinical evaluation if the score was between 6 and 8. The dose could also be decreased, or treatment could be discontinued, if a patient developed persistent side effects. Blood samples were obtained at baseline for DNA extraction, and then again at weeks four and eight for assays of drug and metabolite levels. All patients provided written informed consent. The study protocol was approved by the Mayo Clinic Institutional Review Board.

**Mayo**

The Mayo cohort was collected as a continuation of the PGRN-AMPS cohort. Patients were recruited using the same protocol and inclusion criteria as the PGRN-AMPS cohort described above, but genotyping was carried out separately.

**GENPOD**

A full description of the methodology and sample of the GENetic and clinical Predictors Of treatment response in Depression (GenPod) study can be found elsewhere ^19^. The study was a multi-centre randomized clinical trial of 601 patients with depression (men n=161 women n=347) recruited in primary care who had an ICD-10 diagnosis of major depression of at least moderate severity as assessed by the Clinical Interview Schedule-Revised (CIS-R) ^20^ and the Beck Depression Inventory (BDI) ^21^. Individuals were randomly allocated to either reboxetine (4mg twice daily) or citalopram (20mg). These two drugs represent different mechanisms of antidepressant action with citalopram primarily affecting serotonergic neurotransmission and reboxetine primarily affecting noradrenergic neurotransmission. As ethnicity is a major confounder in genetic studies due to the introduction of LD and haplotypic difference across ethnic backgrounds, only individuals with a white European ancestry were chosen for the whole genome analysis (n=512). Individuals were aged between 18-74 years with a mean age of 38.8 years. Exclusion criteria included if individuals had psychosis, bipolar disorder or major substance or alcohol abuse, or if they had medical contraindications. Individuals were further excluded from the analysis if they had no post baseline information. All participants provided written consent after the study and procedure were explained. Ethical approval was obtained from the South West Ethics Committee (MREC 02/6/076) as well as research governance approval from Bristol, Manchester and Newcastle Primary Care NHS Trusts. The ISRCTN is 31345163 and EudraCT number 2004-001434-16.

**Pfizer**

A total of 345 patients from eight MDD clinical trials were provided. Study designs were variable and primarily conducted as double-blind, placebo-controlled, 6 to 8 weeks studies with sertraline, fluoxetine or paroxetine as active comparators in addition to the investigational compound. In all eight trials, a diagnosis and inclusion criterion for MDD was a HAMD-17 total score of 22 or higher at screening. Exclusion criteria included DSM-IV diagnosis of psychotic features, bipolar I or II, or major risk for suicide. Only subjects from the SRI comparator arms were sent for whole genome genotyping and included in the current study. All study protocols received institutional review board (IRB) approval and informed consent was obtained from participating subjects prior to sample collection.

**GSK**

The samples from GSK derived from two randomized, double-blind, placebo-controlled comparisons of the antidepressant efficacy and the effects on sexual functioning of Bupropion XL and escitalopram in outpatients with moderate to severe depression ^22^. The studies were parallel groups and identically designed, conducted between January 2003 and June 2004 in the United States. Escitalopram or matching placebo capsule was administered at doses of 10mg/day for the first 4 weeks and either 10mg/day, or if clinically indicated, 20mg/day from Week 5 through Week 8. Included subjects had primary diagnosis of MDD with duration at recruitment lasting 12 weeks but no greater than 2 years, and having failed to respond to two adequate trials of antidepressants in the previous 2 years. The primary outcomes for depression was the change from baseline in HAMD-17 total score, whilst the secondary outcomes were percent of subjects in remission and percent of responder (HAMD-17), plus CGI-I and CGI-S. Out of the 210 patients in the escitolapram treatment arms who completed the study, 137 were selected based on availability of consented DNA blood sample and white Caucasian ethnicity; with an average age of 36.4 (from 18 to 64) and 45:55 male to female ratio. All patients provided written informed consent prior to any study activity and the protocol for each of the studies was approved by international review boards.

**GODS**

The Geneva Outpatient Depression Study (GODS) has been described in detail elsewhere ^23,24^. Briefly, GODS is a partly randomized study, which examined the efficacy of four antidepressants (paroxetine, clomipramine, venlafaxine and nefazodone) based on a seven-step algorithm in a cohort of 131 subjects (53 men and 78 women) with severe MDD aged 18 to 65 years. Exclusion criteria included pregnancy, schizophrenia or schizoaffective disorder, dependence on alcohol or other substances and treatment with mood-stabilisers or antipsychotics. The present investigation includes data from the first three steps that included treatment with paroxetine (an SRI), initiated at 20mg daily and increased to 30mg and 40mg daily if remission was not achieved. Patients were discharged only if complete remission was obtained as defined by a MADRS score of 8 or less. Of the 131 GODS participants, 82 had available blood DNA samples and reported white European ancestry. The study protocol was approved by the ethics committee of the Geneva University Department of Psychiatry and written informed consent was obtained from all subjects.

**Miaoli**

Patients within the Miaoli cohort were included within the International SSRI Pharmacogenomics Consortium (ISPC) genome-wide association study (GWAS) of antidepressant response and has been previously described there ^25^. Patients were recruited from outpatient clinics of five hospitals in northern Taiwan. All participants were at least 18 years old and had a depressive episode at their baseline visit with a score of at least 14 on the HAMD-21. Patients were interviewed by board certified psychiatrists and trained research nurses. The clinical diagnosis was made according to DSM-IV criteria, using the Structured Clinical Interview for DSM-IV Axis-I disorders. Those with a primary or a comorbid diagnosis of schizophrenia, schizoaffective disorder, bipolar disorder, alcohol or substance dependence, dementia, or other significant medical conditions, and those who had been treated previously with paroxetine or escitalopram were excluded. Before entering the study, patients needed to have completed a 7-day washout period for any earlier antidepressant treatments (12 days for fluoxetine). Patients were administered either escitalopram or paroxetine according to the judgment of study clinicians. In the escitalopram arm, patients received a daily fixed dose of 10 mg for the first 4 weeks, followed by flexible dosages of 10–30 mg/day on the basis of their clinical response over an 8-week treatment. In the paroxetine arm, patients received a daily fixed dose of 20 mg paroxetine for the first 4 weeks, and then a second 4-week period of flexible dosing (20–40 mg/day). No other psychotropic drugs were allowed during this period, except for 10 mg of zolpidem per night to treat insomnia (as needed, but for no more than four nights per week) and 1–2 mg of lorazepam per day to treat anxiety symptoms as needed. Study participants were assessed using the HAMD-21 and the Hamilton Rating Scale for Anxiety (Hamilton, 1959), at week 0 (baseline or the time of enrollment), 1, 2, 4, 6, and 8 of the continuous treatment period. Additional details about the study can be found in prior publications ^26–28^. This study was approved by the institutional review boards of the National Health Research Institutes and all participating clinics, with the requirement of written informed consent signed by each study participant.

**Taipei**

This sample has been used for a number of candidate gene pharmacogenetic studies ^29–31^, and was part of the International SSRI Pharmacogenomics Consortium (ISPC) genome wide association study ^25^. Briefly, the case population was comprised of patients diagnosed with a current major depressive episode fulfilling DSM-IV criteria. Each diagnosis was made by one board-certificated and experienced psychiatrist in accordance with DSM-IV. Additional inclusion criteria were a minimum score of 18 on the 21-item HAMD, presence of depressive symptoms, and antidepressant-naive/free for at least 2 weeks prior to study commencement. Exclusion criteria were extra DSM-IV Axis I diagnoses, personality disorders, pregnancy, recent suicide attempt, and major medical and/or neurological disorders. The sample consisted entirely of ethnically Chinese adults. For the pharmacogenetic study, daily doses of fluoxetine or citalopram were given, starting at 20 mg/day; based on the clinical response the investigator could increase the dosage to 40 mg/day. No other psychotropic medications were permitted; however, anxiolytics were allowed for insomnia. Treatment efficacy was evaluated by the same investigator, blind to patient genotype, who administered the HAMD prior to treatment, in the 1st week, 4th week and 8th week.

**Japan**

Patients included in this study participated in randomized controlled trials of antidepressants at the Kansai Medical University, Osaka ^32–34^, and were part of the International SSRI Pharmacogenomics Consortium (ISPC) genome wide association study ^25^. Diagnosis of major depression was confirmed by the Structured Clinical Interview for DSM-IV Axis I Disorders (SCID-I). Exclusion criteria were as follows: clinically significant unstable medical illness, pregnancy, main psychiatric diagnosis other than major depression (including dementia, mental retardation, substance abuse, dysthymia, panic disorder, obsessive–compulsive disorder and generalized anxiety disorder), and electroconvulsive therapy within the previous 6 months. The diagnoses were made by two independent senior psychiatrists and confirmed by a third psychiatrist. All patients were evaluated at baseline and bi-weekly thereafter until week 6 using the 21-item HAMD administered by a trained senior psychiatrists blind to genetic data. Patients were either drug free or taking ineffective antidepressants and after ten days of washout, paroxetine (n = 68) or fluvoxamine (n = 54) was administered to reach therapeutic doses from days 8 to 11 until the end of trial (fluvoxamine: 150 mg/day; paroxetine: 40 mg/day). The type of prescribed SSRI remained unchanged throughout the study period. The study was approved by the ethical committee of the Kansai Medical University and Osaka University. Written informed consent was obtained from each subject after a detailed explanation of the study.

#### Phenotype definitions

Two measures of antidepressant response were defined, Remission and Percentage Improvement. Remission is a binary measure attained when a patient’s depression symptom score decreases to a pre-specified threshold for the rating scale. Remission thresholds for scales used in these studies were: MADRS ≤ 10, QIDCS ≤ 5, HAMD-17 ≤ 7, HAMD-21 ≤ 7, and BDI ≤ 9. Patients who did not reach these thresholds were classified as non-remitting. The quantitative measure of Percentage Improvement was calculated as 100*(baseline score – final score)/baseline score. Thus, a higher Percentage Improvement implies a better treatment response and a negative Percentage Improvement implies the patient’s symptom score worsened during treatment. Percentage Improvement was standardised into a Z-score within each cohort.

#### Quality control and imputation of genotype data using RICOPILI

Genotyping procedures can be found in the primary reports for each cohort (summarized in Supplementary Table 1). Individual genotype data for all cohorts were processed using the Psychiatric Genomics Consortium (PGC) “RICOPILI” pipeline for standardized quality control, imputation, and association analysis ^35^. Imputation of SNPs and insertion-deletion polymorphisms was performed using the 1000 Genomes Project multi-ancestry reference panel ^36^.

Quality control and imputation were performed separately for European and East Asian cohorts according to standards from the PGC. The ancestry of samples is determined by comparison to the 1000 Genome reference and ancestral outliers are removed using EIGENSTRAT ^37^, thereby ensuring samples from different cohorts are within the same ancestral cluster. The default parameters for retaining SNPs and subjects were: SNP missingness < 0.05 (before sample removal); subject missingness < 0.02; autosomal heterozygosity deviation (|*F_het_*|<0.2); SNP missingness < 0.02 (after sample removal); difference in SNP missingness between remitters and non-remitters < 0.02; and SNP Hardy-Weinberg equilibrium (*P*>10^−6^ in remitters or *P*>10^−10^ in non-remitters). These default parameters sufficiently controlled λ and false positive findings for all cohorts.

Genotype imputation was performed using the pre-phasing/imputation stepwise approach implemented in IMPUTE2 / SHAPEIT (chunk size of 3 Mb and default parameters). The imputation reference set consisted of 2,186 phased haplotypes from the 1000 Genomes Project dataset (August 2012, 30,069,288 variants, release “v3.macGT1”). After imputation, we identified SNPs with very high imputation quality (INFO >0.8) and low missingness (<1%) for building the principal components to be used as covariates in final association analysis. Principal components were derived within each cohort separately. SNPs underwent linkage disequilibrium-based pruning (r^2^ > 0.02) and frequency filtering (MAF > 0.05). This SNP set was used for robust relatedness testing and population structure analysis. Relatedness testing identified pairs of subjects with $\hat{\pi}$ > 0.2, and one member of each pair was removed at random after preferentially retaining remitters over non-remitters. Principal component estimation used the same collection of autosomal SNPs.

#### Genome-wide association analysis

GWAS was performed using the RICOPILI pipeline, which tests for association between phenotype and genotype dosages within each sample with logistic regression (for Remission) and linear regression (for Percentage Improvement) ^35^. Genome-wide results from each study were meta-analysed using inverse-variance weighting with METAL ^38^. GWAS meta-analysis was performed separately for studies with participants of European and of East Asian ancestry. Meta-analysis across these ancestral groups was not performed though a comparison of results was performed (described below in the ‘Comparison between East Asian and European antidepressant GWAS’ section). A genome-wide significance threshold of 5×10^-8^ was used ^39^.

#### Estimation of SNP-heritability

The SNP-heritability of Remission and Percentage Improvement was estimated using individual-level data by GREML (genomic-relatedness-based restricted maximum-likelihood) in GCTA (Genome-wide Complex Trait Analysis) software ^40,41^. GREML was performed after merging genotype data across studies to construct a single genomic relatedness matrix (GRM) based on all overlapping genetic variants with frequency of >1% and missingness of <1% in all studies. The GRM was adjusted for incomplete tagging of causal variants using GCTA, and in addition to the relatedness checks performed by RICOPILI, with 210 individuals were removed by GCTA based on a relatedness threshold of >0.05. The number of individuals remaining in the GREML analysis was 5,117 for Percentage Improvement, and 5,051 for Remission (N_remitters_= 1,805; N_non-remitters_= 3,246). The heritability from GREML was not constrained to >0 and <1. The analysis was performed across all cohorts simultaneously (mega-GREML), and separately within each cohort and then inverse variance meta-analysed (meta-GREML). Comparison of mega- and meta-GREML estimates can provide insight into the heterogeneity between cohorts.

To verify GREML SNP-heritability estimates, two methods with distinct assumptions were also used; GCTB ^42^ and Linkage Disequilibrium-Score Regression (LDSC) ^43^. Due to sample size requirements of these methods, they were only used to estimate the SNP-heritability across samples, equivalent to mega-GREML. GCTB’s nested Bayes-S model with default settings was applied to the individual-level genotype data. LDSC was performed using the GWAS summary statistics, using the LDSC munge_sumstats.py script with default settings. The SNP-heritability was estimated with and without constraint of the intercept.

SNP-heritability estimates for Remission were transformed to the lability scale to enable comparison of SNP-heritability estimates between cohorts and with future studies, using a population prevalence of 30% for remission. However, given this study ascertained individuals based on a diagnosis with depression, and the liability model is based on population theory, using a liability conversion assumes that there is no correlation between liability for depression and the likelihood of remission after antidepressant treatment.

#### Leave-one-out polygenic scoring

To determine whether the Remission and Percentage Improvement GWAS within core samples could be used to predict antidepressant response in an independent sample, a leave-one-out polygenic scoring approach was employed. This involves calculating polygenic scores within each cohort based on GWAS summary statistics derived using all other cohorts. Polygenic scores were calculated using default settings in PRSice V2 ^44^. PRSice performs linkage disequilibrium (LD)-based clumping to account for LD and then calculates multiple polygenic scores based on SNPs achieving the default 8 *p*-value thresholds when using the --fastscore option. The European 1000 Genomes Phase 3 reference was used to estimate LD between variants for clumping. Covariates were regressed from the polygenic scores within each cohort, and the association between the PRS residuals and antidepressant response measures was then assessed across all cohorts simultaneously. One-sided p-values were used to assess statistical significance as this analysis.

#### PRS replication cohorts

**Janssen**

Janssen data was drawn from a Janssen clinical study termed ‘RIS-INT-93’ (ClinicalTrials.gov number [NCT00044681](about:blank))^45^. The study includes participants meeting the DSM-IV criteria for MDD and had history of resistance to antidepressant therapy, defined as failure to respond to at least one but not more than three adequate antidepressant trials during the current episode. Open-label prospective treatment of individuals with citalopram for up to 6 weeks was carried out. Depressive symptoms were assessed using the HAMD-17. All subjects signed written informed consent before participation, with the protocol approved by institutional review boards at participating institutions.

**Douglas Biomarker Study**

The Douglas Biomarker Study involved an 8-week antidepressant treatment for MDD patients randomly selected to receive either densvenlafaxine or escitalopram^46^. HAMD scores were used to assess symptom severity at baseline and week 8. Participants were recruited at the Depressive Disorders Program at the Douglas Mental Health Institute, McGill University (Montreal, QC).

**IRL-GREY**

IRL-GREY, the ‘Incomplete Response in Late Life Depression: Getting to Remission’ study^47^, is a randomised placebo-controlled clinical trial conducted to test the efficacy, safety, and tolerability of aripiprazole augmentation in older adults whose depression had not remitted with venlafaxine. Participants were aged ≥60 with a current SCID or DSM-IV diagnosis of MDD and a MADRS score of ≥15. All participants provided written, informed consent and the study was overseen by a Data Safety and Monitoring Board.

**Generation Scotland**

Generation Scotland is a family and population-based cohort in Scotland linked to prescribing records. Of the 20,032 individuals with genetic data, 3,452 (17.2%) individuals were prescribed at least one antidepressant. Individuals within this subset were classified as having treatment resistant depression (TRD) if they switched antidepressants two or more times(antidepressant switching was considered as a proxy of non-response) ^48^. Analyses in Generation Scotland were controlled for age, sex, and 20 principal components of population structure.

**AGDS**

AGDS is a population-based cohort in Australia of individuals reporting a diagnosis of major depressive disorder ^49^. Participants were asked to report on the efficacy of antidepressants, with 11,247 genotyped individuals providing information on their use of SSRIs. Participants were defined as responders if they reported ‘responding very well’ (N=4,368), and non-responders if they report ‘only moderate or poor response’ (N=6,879). Analyses in AGDS were controlled for 10 principal components of population structure.

#### Gene-level association analysis

Gene associations were estimated using MAGMA (Multi-marker Analysis of GenoMic Annotation)^50^ and TWAS (Transcriptome-Wide Association Study)^51^.

MAGMA v1.06b SNP-wise mean model (±10kb window) was used to perform gene-level association analysis based on the Remission and Percentage Improvement GWAS p-values. The analysis was based on genetic variants available in the 1000 Genomes Phase 3 dataset available on the MAGMA website (‘g1000_eur.bim’). SNPs were assigned to genes using the MAGMA ‘NCBI37.3.gene.loc’ file with a 10kb window. The LD between variants was based on the MAGMA release of the 1000 Genomes Phase 3 reference (‘g1000_eur.bed/.bim/.fam’). False Discovery Rate (FDR) correction was used to control for multiple testing.

TWAS integrates GWAS associations with external eQTL data to infer whether differential gene expression estimated from SNP data is associated with the GWAS phenotype. TWAS was performed using FUSION software ([http://gusevlab.org/projects/fusion/](about:blank)), and precomputed multi-SNP predictors of gene expression based on data collected from multiple specific brain regions, thyroid tissue, pituitary gland, liver and blood (Table S3). The transcriptome-wide significance threshold of *p*<2.51x10^-6^ was estimated using a permutation procedure ^52^. To test whether the same causal SNP affects both the GWAS phenotype and gene expression, colocalization analysis was performed using the ‘coloc’ package in R ^53^, as implemented by FUSION software.

#### Gene-set and tissue enrichment analysis

Enrichment of genes associated with antidepressant response in gene-sets and tissues was evaluated using MAGMA. Two groups of gene-sets were evaluated: 1) A previously curated list of 76 gene-sets previously implicated in psychiatric phenotypes ^54^, and 2) 1,751 gene-sets defined by drug interactions collated by DrugTargetor ^55^. Tissue enrichment was performed using data derived from the Genotype-Tissue Expression (GTEx) project ^56^.

To define significant associations, MAGMA’s permutation procedure was used to account for multiple testing within each of these two gene-set enrichment analyses. For the drug interaction gene set analysis, we used a one-way Wilcoxon–Mann–Whitney test to determine post-hoc whether antidepressant or antipsychotic medications were enriched.

Tissue enrichment was performed using the gene property analysis MAGMA v1.06b and the gene-level associations derived using MAGMA described above. Median gene expression levels for each gene and tissue was downloaded from the GTeX V7 consortium (https://gtexportal.org/home/datasets). Genes with a TPM >1 in at least one tissue were retained, and TPM values greater than 50 were windsorized, and then log transformed with a pseudo count 1, in accordance with the FUMA pre-processing (https://fuma.ctglab.nl/tutorial#magma). To derive a measure of preferential expression for each gene, the expression level of each gene within each tissue was then standardized based on the mean and standard deviation of expression across all tissues. Acknowledging the limited power of the Remission and Percentage Improvement GWASs, analysis was initially restricted to test for enrichment of genes expressed in the brain, or HPA-axis tissues. Tissue-specific enrichment was subsequently performed to show the contribution of each tissue to the brain and HPA-axis enrichment results. Comparison between East Asian and European antidepressant GWAS

No statistically significant enrichment of tissues or candidate gene sets were identified for either Remission or Percentage Improvement. Full results are shown in Figure S12 and Tables S20-S21. Similarly, no drug-binding gene-sets were significantly enriched. Looking across drug-binding gene-set results using the one-way Wilcoxon-Mann-Whitney test showed a nominally significant enrichment of antipsychotic gene-sets for Percentage Improvement (*p*=0.020), but not Remission (*p*=0.99). Antidepressant gene-sets showed no evidence of enrichment for Remission (*p*=0.99) or Percentage Improvement (*p*=0.31).

#### Comparison of European and East Asian GWAS results

Although the East Asian sample size was too low for discovery of genome-wide significant loci (For QQ-plots see Figures S13-14), replication of suggestive loci (p< 1e-5) from the European GWAS is of interest. We investigated this using two approaches.

First, we retrieved East Asian GWAS results for the lead SNPs for suggestive loci in the European GWAS. We did not use an LD-based proxy approach, as the differences in LD and minor allele frequency make this approach somewhat in appropriate. For remission, 14 of the 18 European lead SNPs were available in East Asian samples, 7 of which had the same direction of effect, and none of which achieved nominal significance. For remission, 10 of the 18 European lead SNPs were available in East Asian samples, 6 of which had the same direction of effect, and one of which achieved nominal significance (rs570948877, p-value = 0.05). rs570948877 is nearest (8.9kb upstream) to the *SLITRK1* gene encoding a transmembrane and signalling protein that is part of the SLITRK gene family, which is responsible for synapse regulation and presynaptic differentiation in the brain ^57^.

The second approach was to check whether any loci achieving suggestive significance in the European GWAS, also contained evidence of association in the East Asian GWAS (Tables S7-S8). Here we weren’t matching lead SNPs but instead checking whether any SNP within a 250kb window of the European lead SNP achieved a p-value < 0.001. No European suggestive loci for Remission contained evidence of association in the East Asian Remission GWAS. Two European suggestive loci for Percentage Improvement showed some evidence of association, and locus plots were generated to visualise this (Supplementary Figures 15 and 16). The lead European SNP for these two loci are rs2080632 and rs73001560, for which the nearest genes can be found in Table S8.

Caution when interpreting these loci should be taken as these replications are not statistically robust due to unaccounted for multiple testing of loci or SNPs within each locus.

#### MAGMA gene association results

MAGMA identified a significant association on chromosome 17 for the gene *ETV4* with both Remission (*p.fdr*=0.016) and Percentage Improvement (*p.fdr*=0.016). Within the same region, *DHX8* was also significantly associated with Remission (*p.fdr*=0.046). The SNP-associations within this region span multiple genes (Figure S17). Full MAGMA gene-based association results in Tables S16-S17.

#### Transcriptome-wide Association Study (TWAS) Results

The GWAS summary statistics contained sufficient signal to impute 58,517 features for Remission, consisting of 14,529 unique genes, and 59,236 features for Percentage Improvement, consisting of 14,714 unique genes. TWAS identified no association achieving transcriptome-wide significance (*p*<2.51x10^-6^). Further inspection of TWAS associations within the chromosome 17 region implicated by MAGMA highlighted SNP-associations with upregulation of *BRCA1* (Remission *p*=1.96x10^-4^; Percentage Improvement *p*=9.21x10^-5^; GTeX Brain Caudate basal ganglia) and upregulation of *TMEM106A* (Remission *p*=0.0011; Percentage Improvement *p*=0.0018; YFS Blood). Colocalisation analysis of these associations indicated shared casual variants for these gene’s differential expression and antidepressant response. Full TWAS results are given in Supplementary Table 18 and 19.

#### SNP-based heritability sensitivity analysis

Mega-GREML SNP-based heritability estimates were congruent with those estimated using Genome-wide Complex Trait Bayesian Analysis (GCTB) and LDSC (Figure S18; Tables S6 and S9).

Due to sample size limitations, within-sample heritability could not be estimated for all samples. However, mega-GREML results were consistent when excluding these samples (Remission *h*^2^=0.136, SE=0.060, liability scale assuming population prevalence of 0.3; Percentage Improvement *h*^2^=0.017, SE=0.044), indicating the mega- and meta-SNP-based heritability estimates are comparable.

Using mega-GREML, the SNP-based heritability for Remission was estimated on a liability scale assuming a range of population prevalence. Results show that SNP-heritability for Remission is significantly different from zero regardless of the population prevalence assumed (Figure S23 and Table S22). The SNP heritability does not change substantially for prevalence of between 0.2 and 0.4, and the heritability is above 0.1 for values of population prevalence above 0.1.

There is a small discrepancy between the samples considered for Remission and Percentage Improvement due some individuals dropping out during the contributing studies. To determine whether these dropouts are responsible for the apparent difference in SNP-based heritability between Remission and Percentage Improvement, SNP-based heritability was also estimated using only individuals with data available for both measures of antidepressant response (N=4,935). Mega-GREML estimates of SNP-based heritability were highly concordant when restricting the analysis to the 4,935 individuals with data for both Remission and Percentage Improvement (Remission *h*^2^=0.136; SE=0.057; liability scale assuming population prevalence of 0.3; Percentage Improvement *h*^2^=-0.016; SE=0.034).

### Supplementary Figures

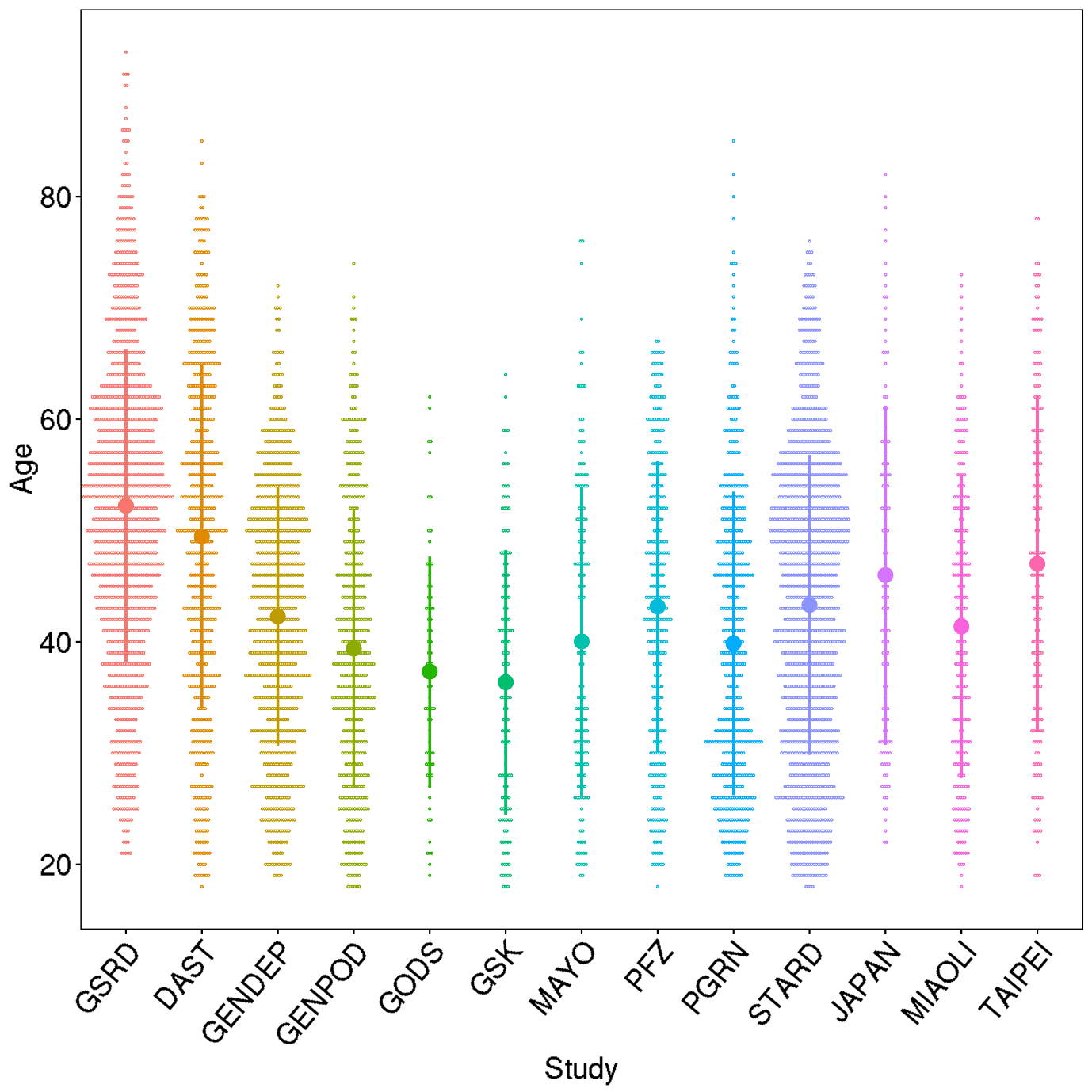

Figure 1. Age across cohorts

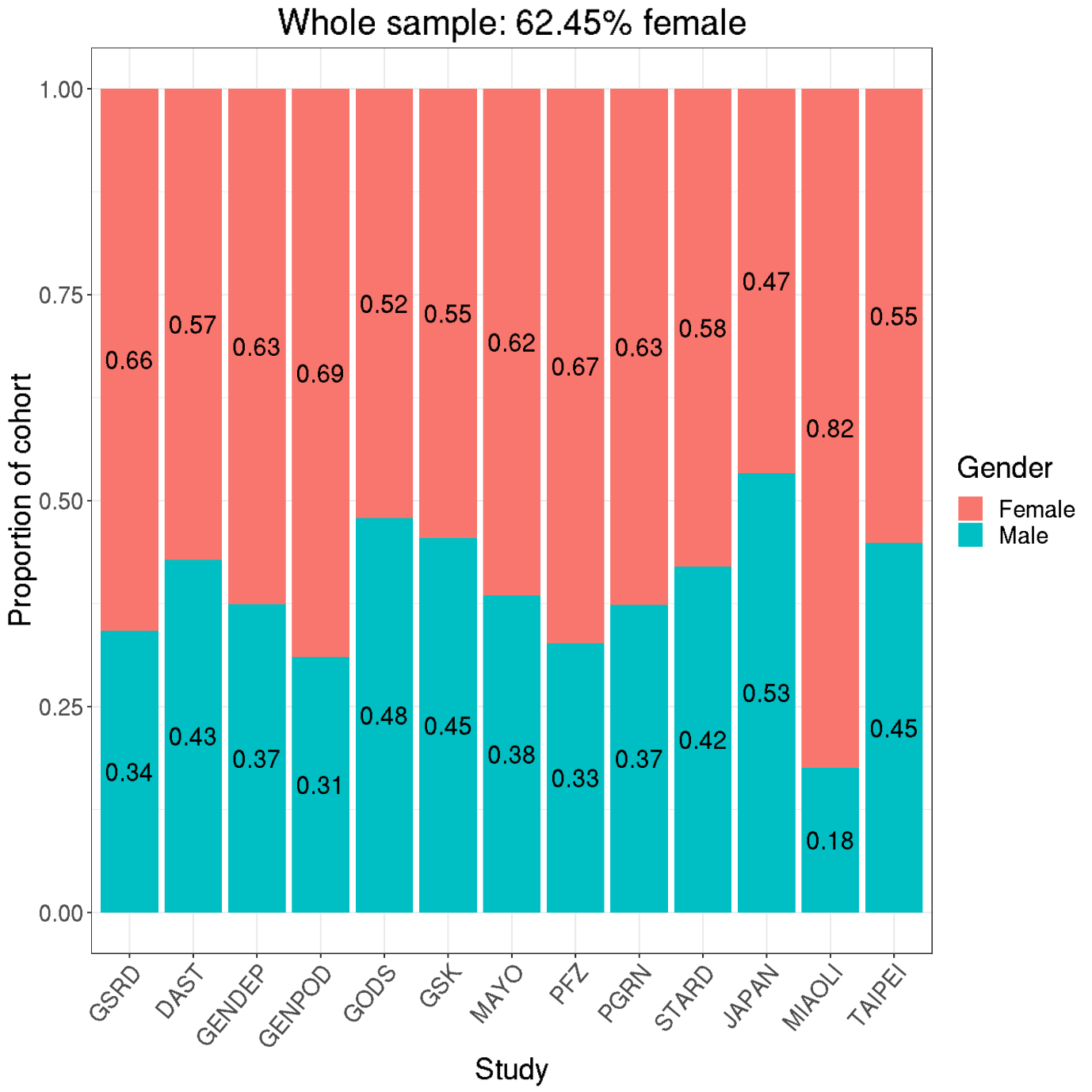

Figure 2. Female ratio across samples

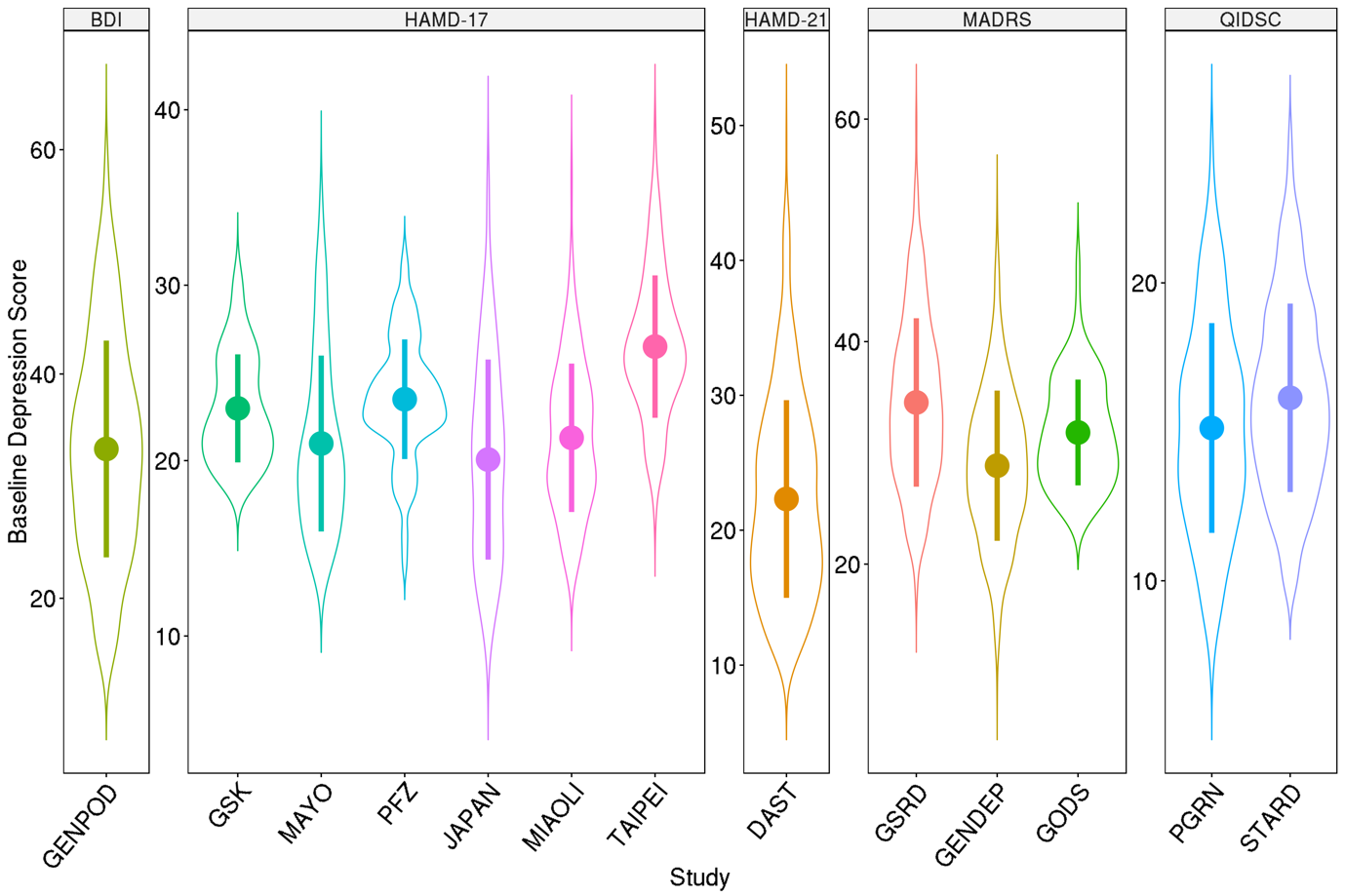

Figure 3. Baseline depression severity across cohorts

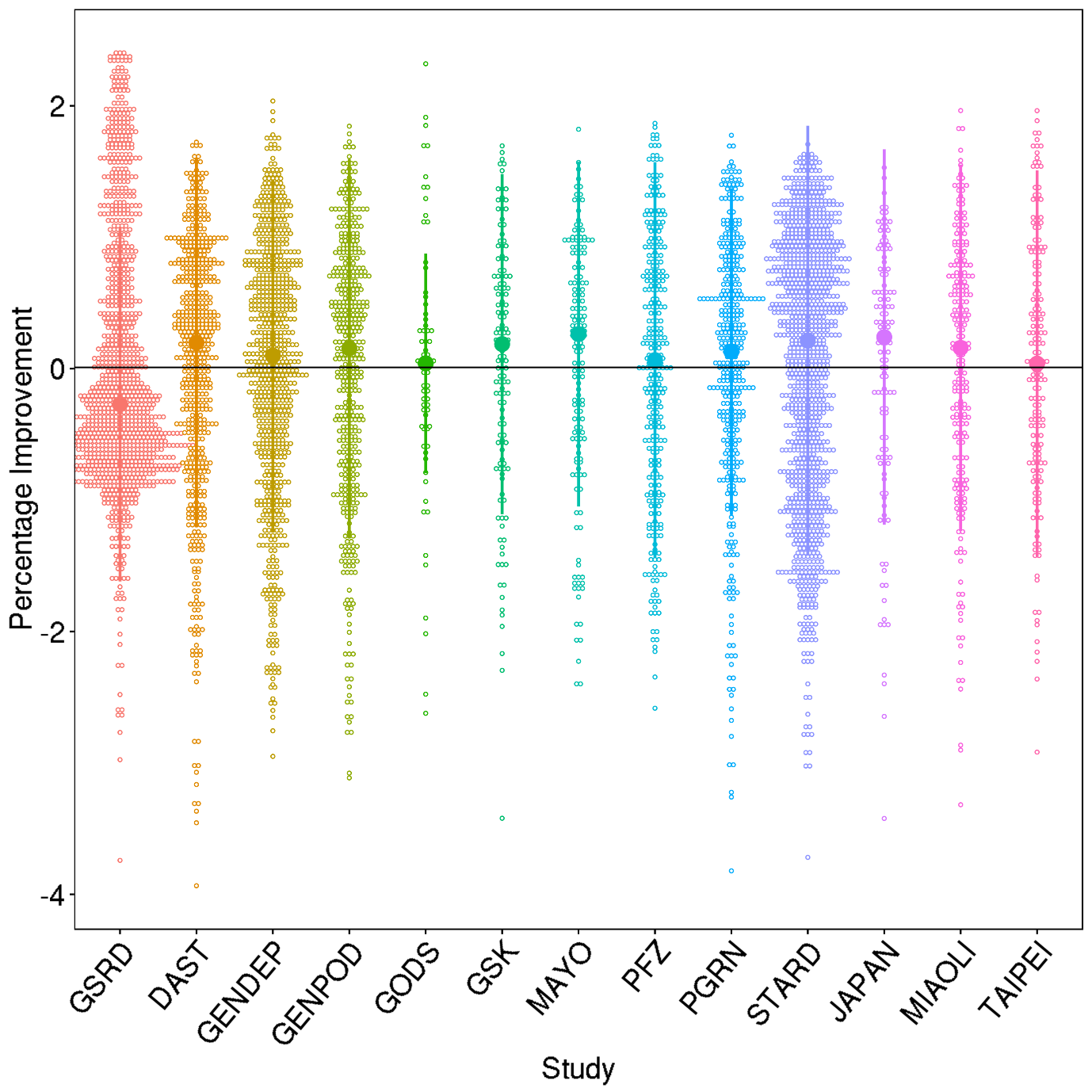

Figure 4. Distribution of percentage improvement after regressing out age, sex, and baseline severity.

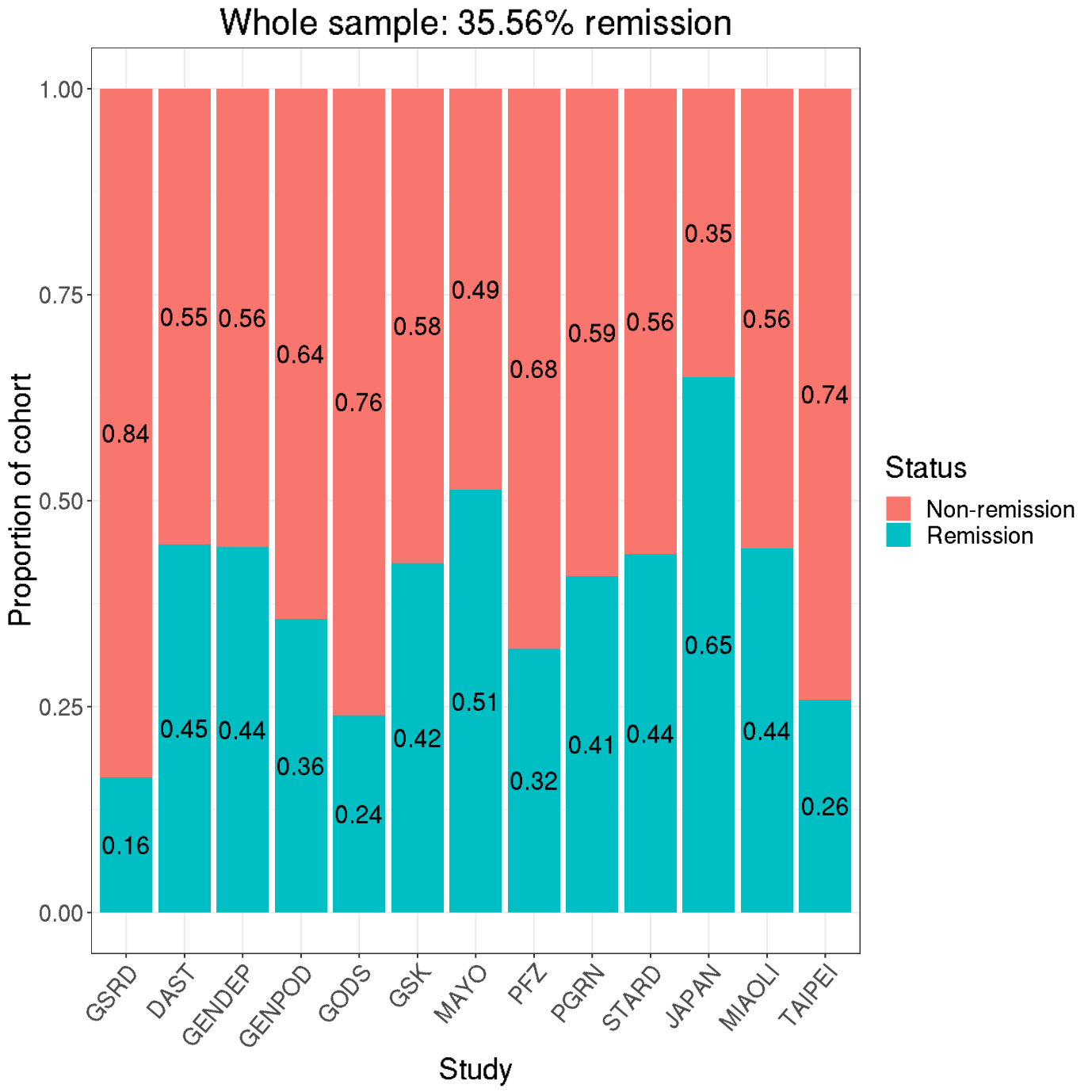

Figure 5. Proportion of sample remitting across cohorts.

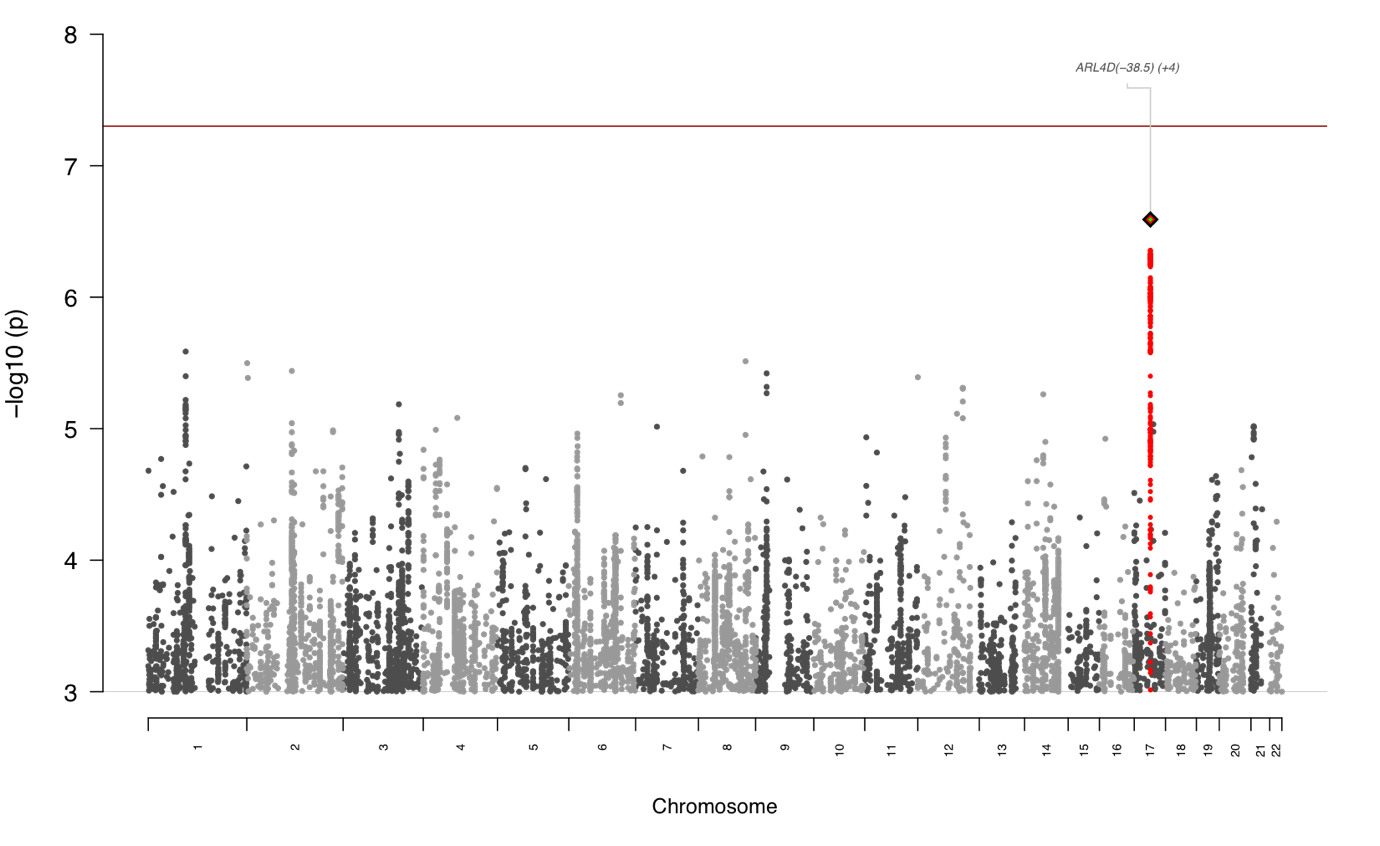

Figure 6. Manhattan plot for European Remission GWAS.

*Note. Showing only variants with -log_10_(p) > 3.*

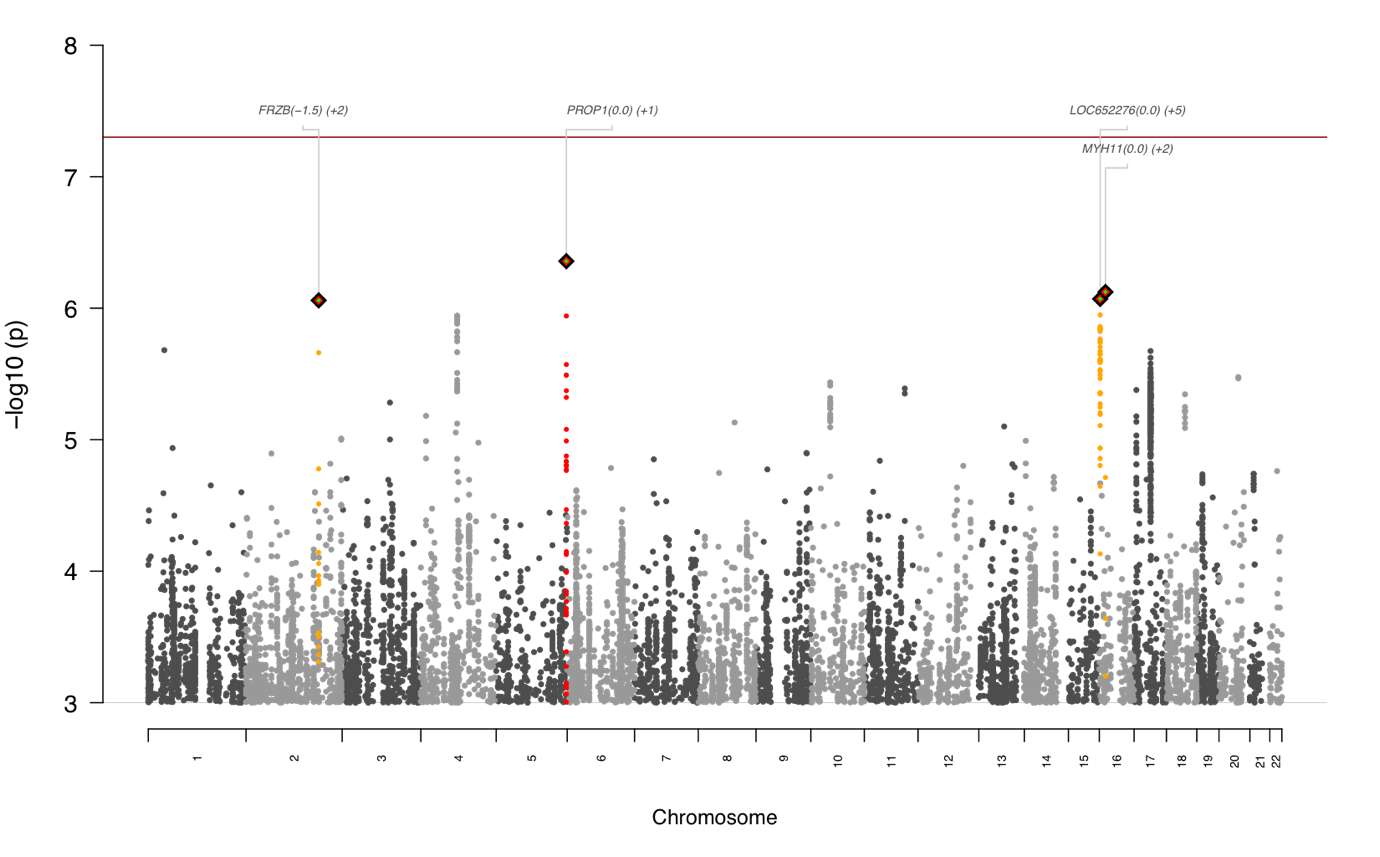

Figure 7. Manhattan plot for European Percentage Improvement GWAS.

*Note. Showing only variants with -log_10_(p) > 3.*

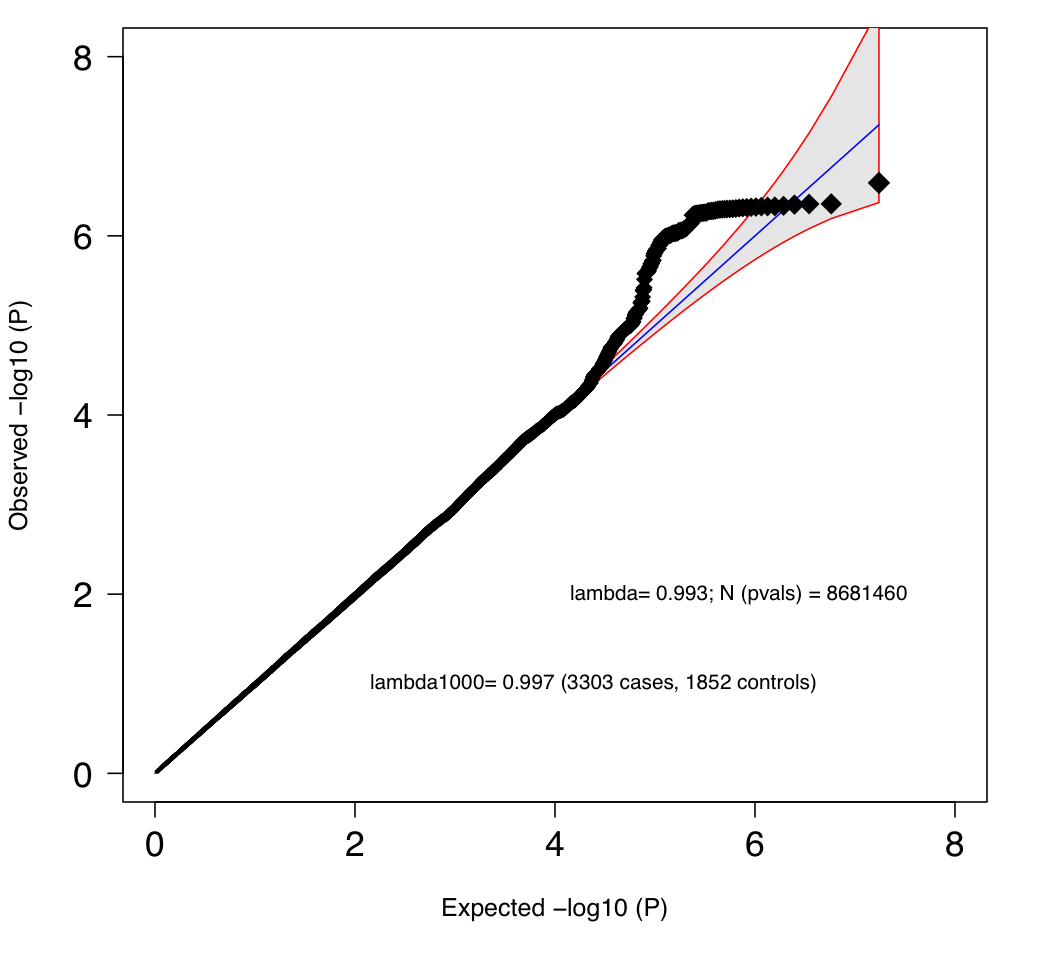

Figure 8. QQ-plot for European Remission GWAS

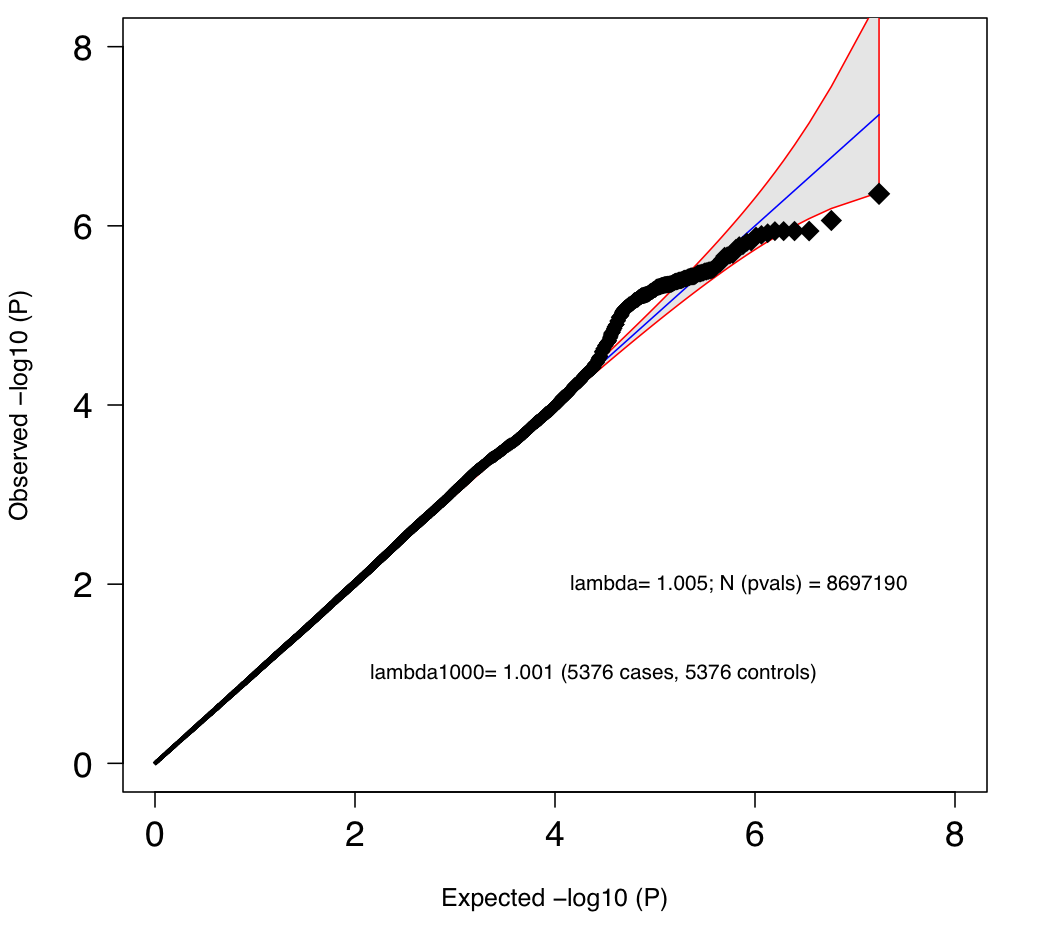

participants)

Figure 9. QQ-plot for European Percentage Improvement GWAS

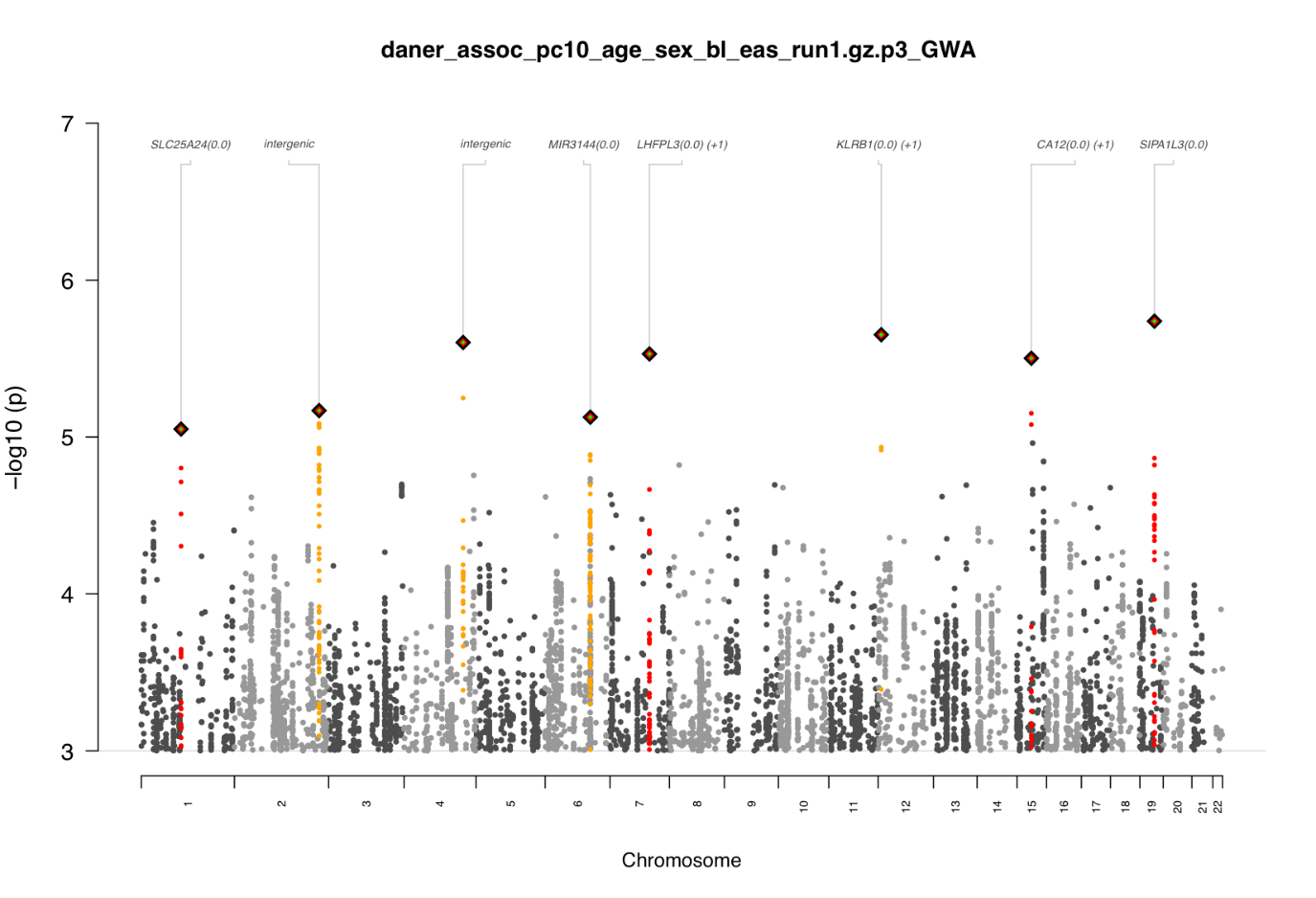

Figure 10. Manhattan plot for East Asian GWAS of Remission.

*Note. Showing only variants with -log_10_(p) > 3.*

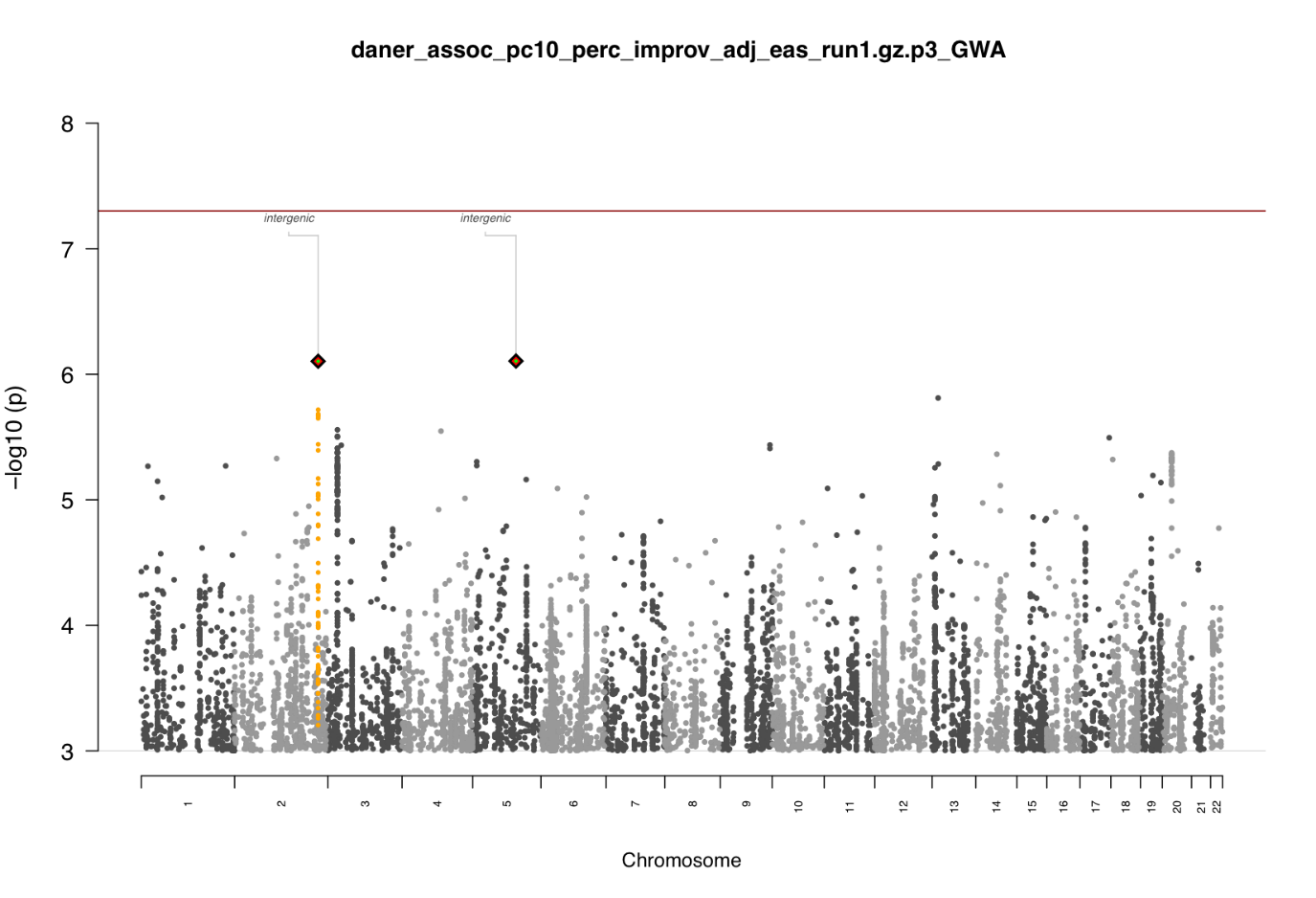

Figure 11. Manhattan plot for East Asian GWAS of Percentage Improvement.

*Note. Showing only variants with -log_10_(p) > 3.*

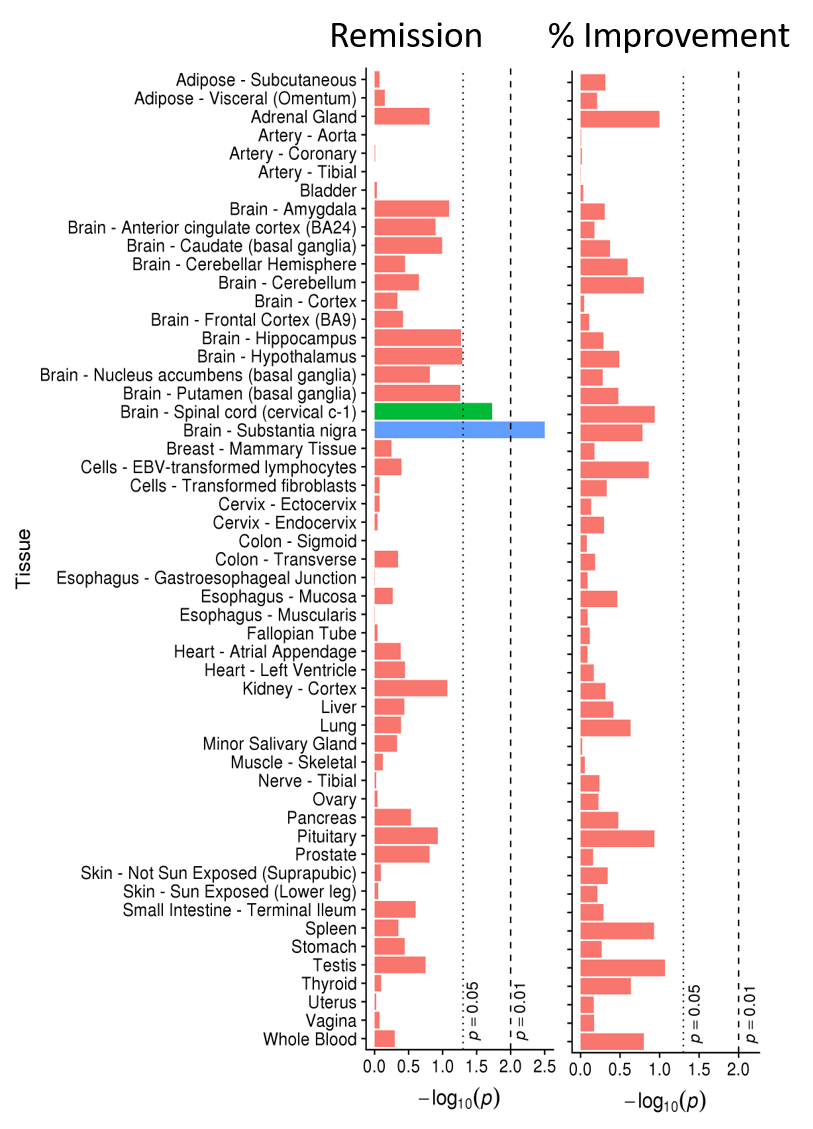

Figure 12. Tissue enrichment results for Remission and Percentage Improvement.

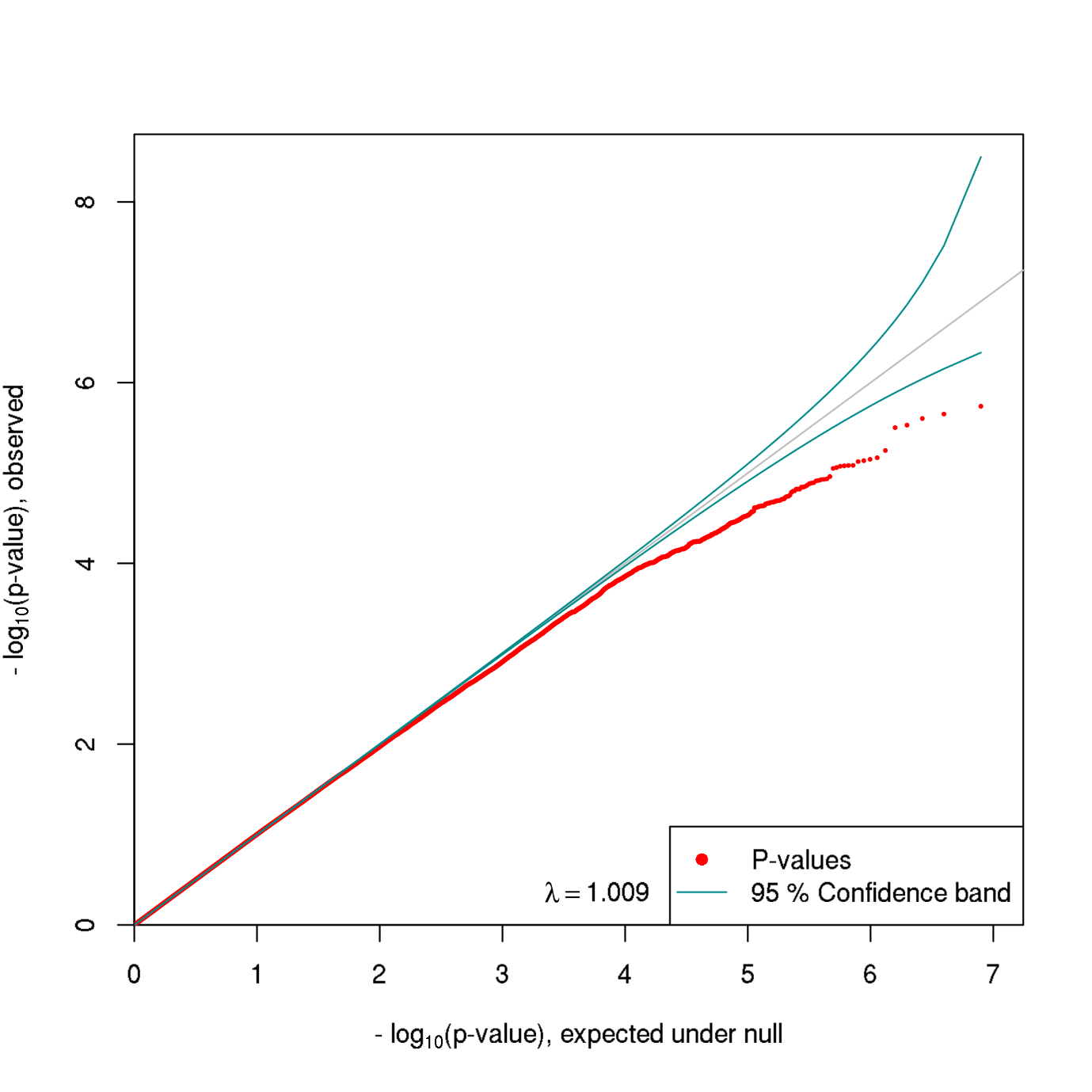

Figure 13. QQ-plot for East Asian Remission GWAS

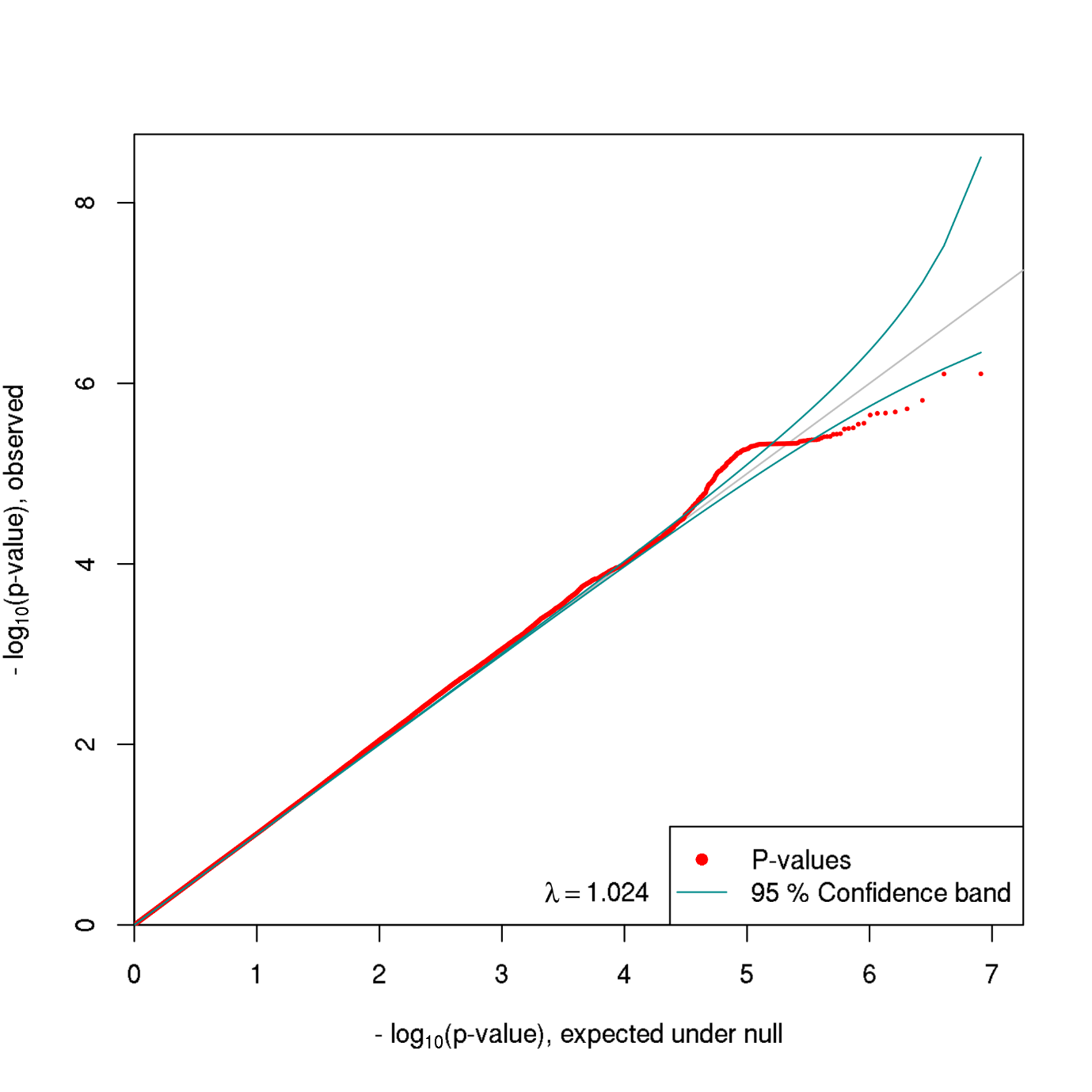

Figure 14. QQ-plot for East Asian Percentage Improvement GWAS

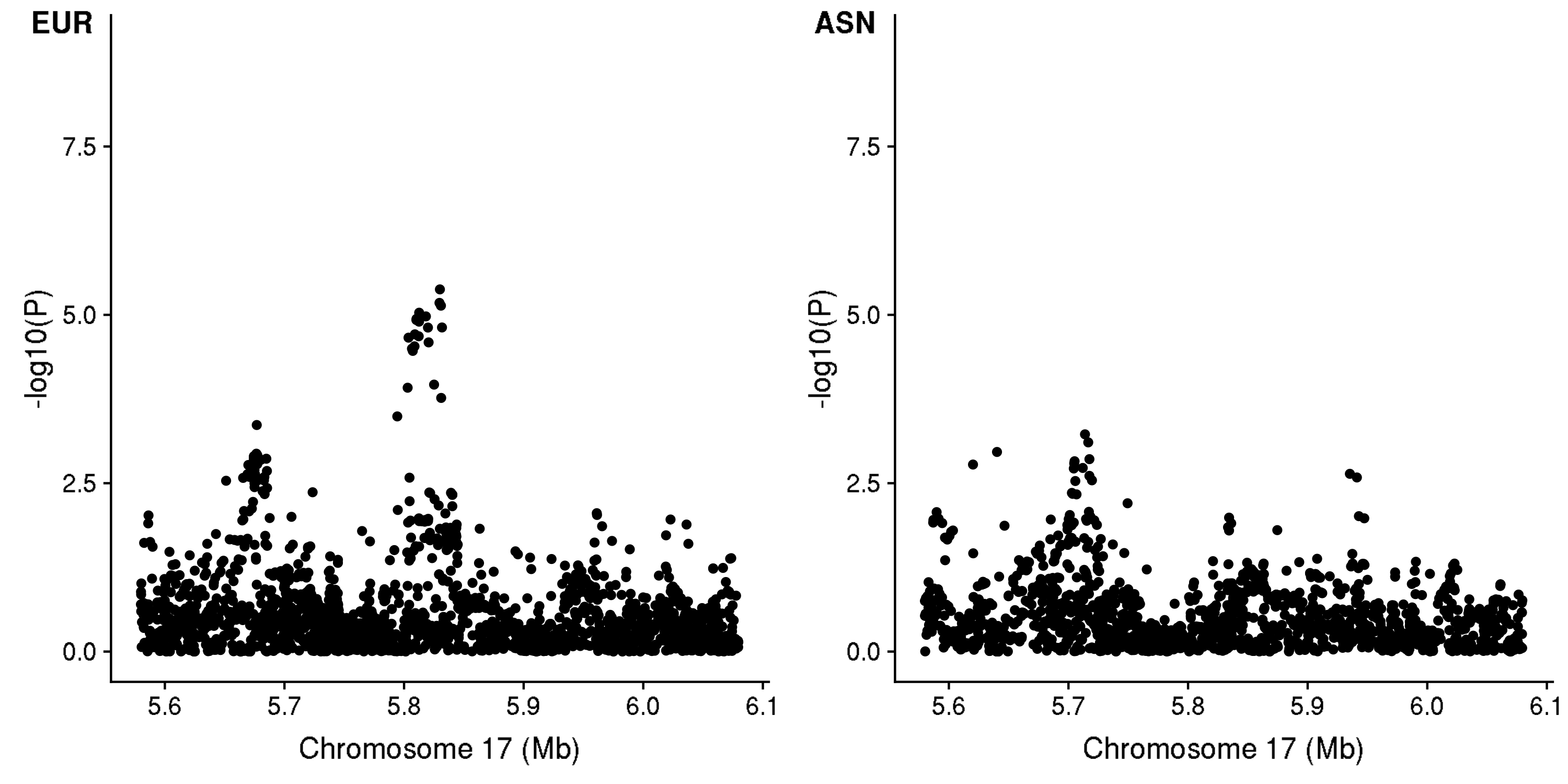

Figure 15. Locus plot of overlapping region of association for Percentage Improvement.

*Note. The European lead SNP of this locus is rs2080632.*

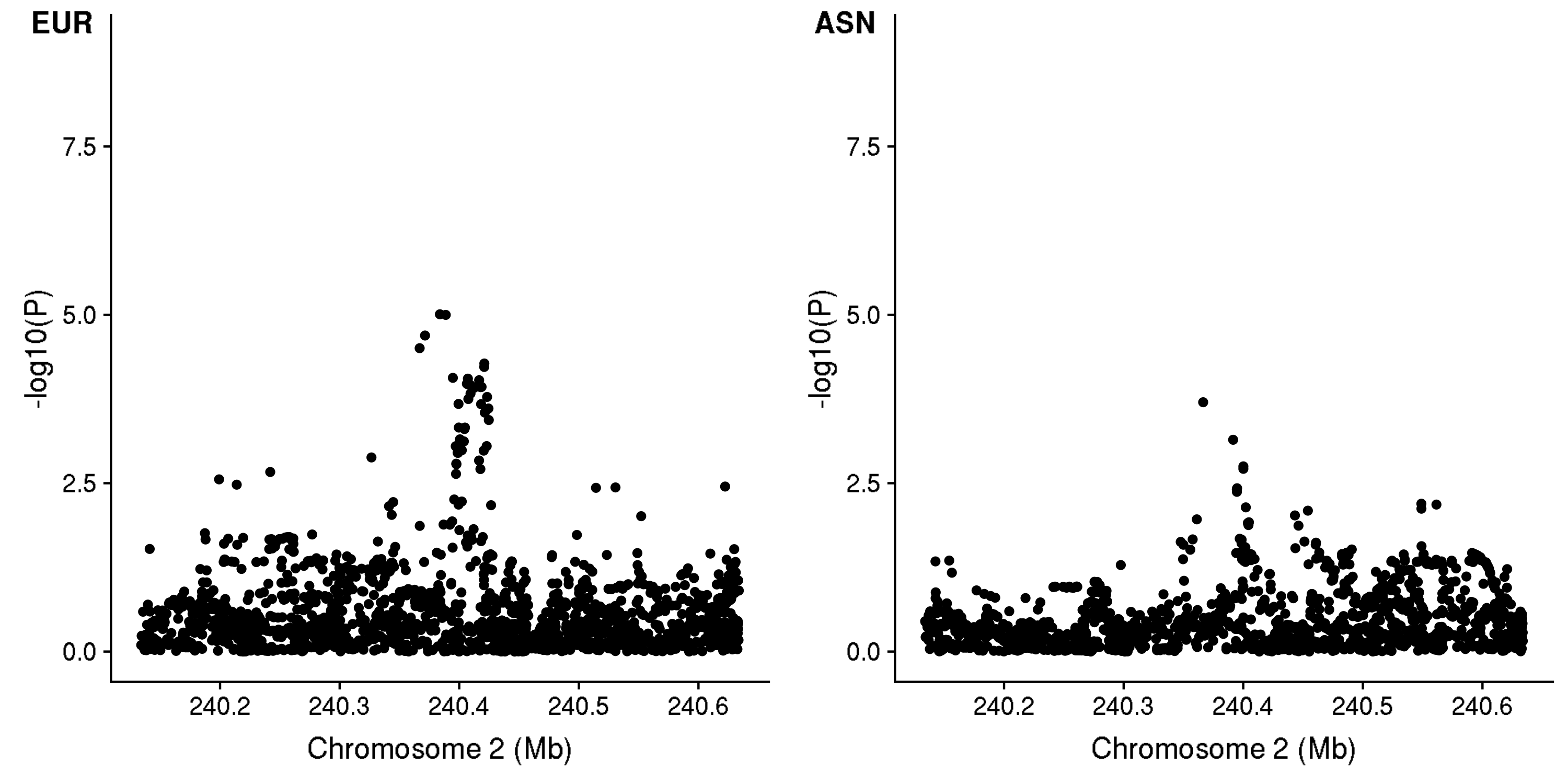

Figure 16. Locus plot of overlapping region of association for Percentage Improvement.

*Note. The European lead SNP of this locus is rs73001560.*

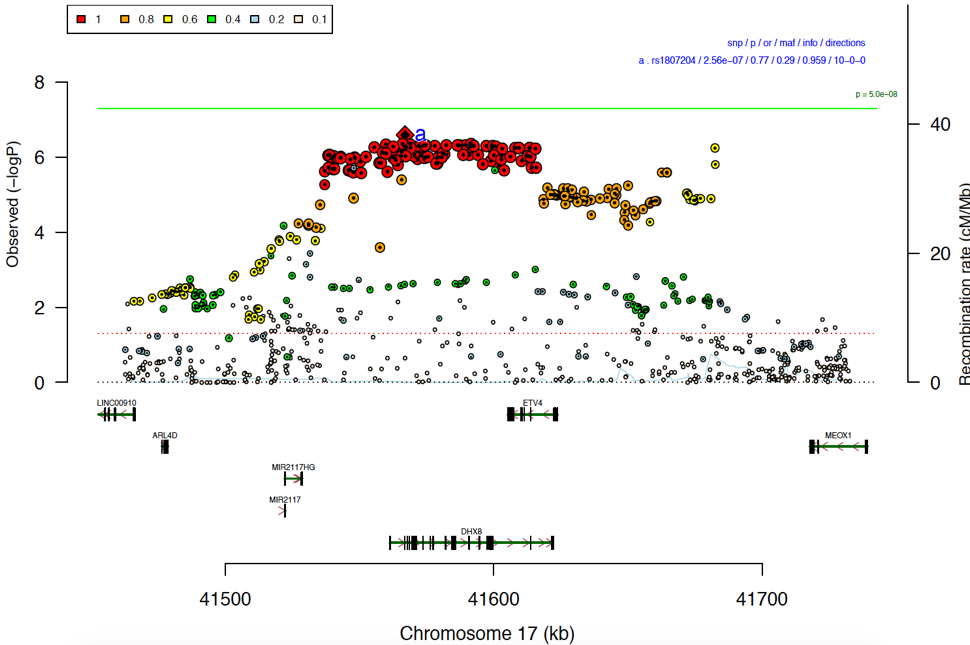

Figure 17. Locus plot showing SNP associations for Remission within the locus containing the significant gene-based association with ETV4 identified by MAGMA.

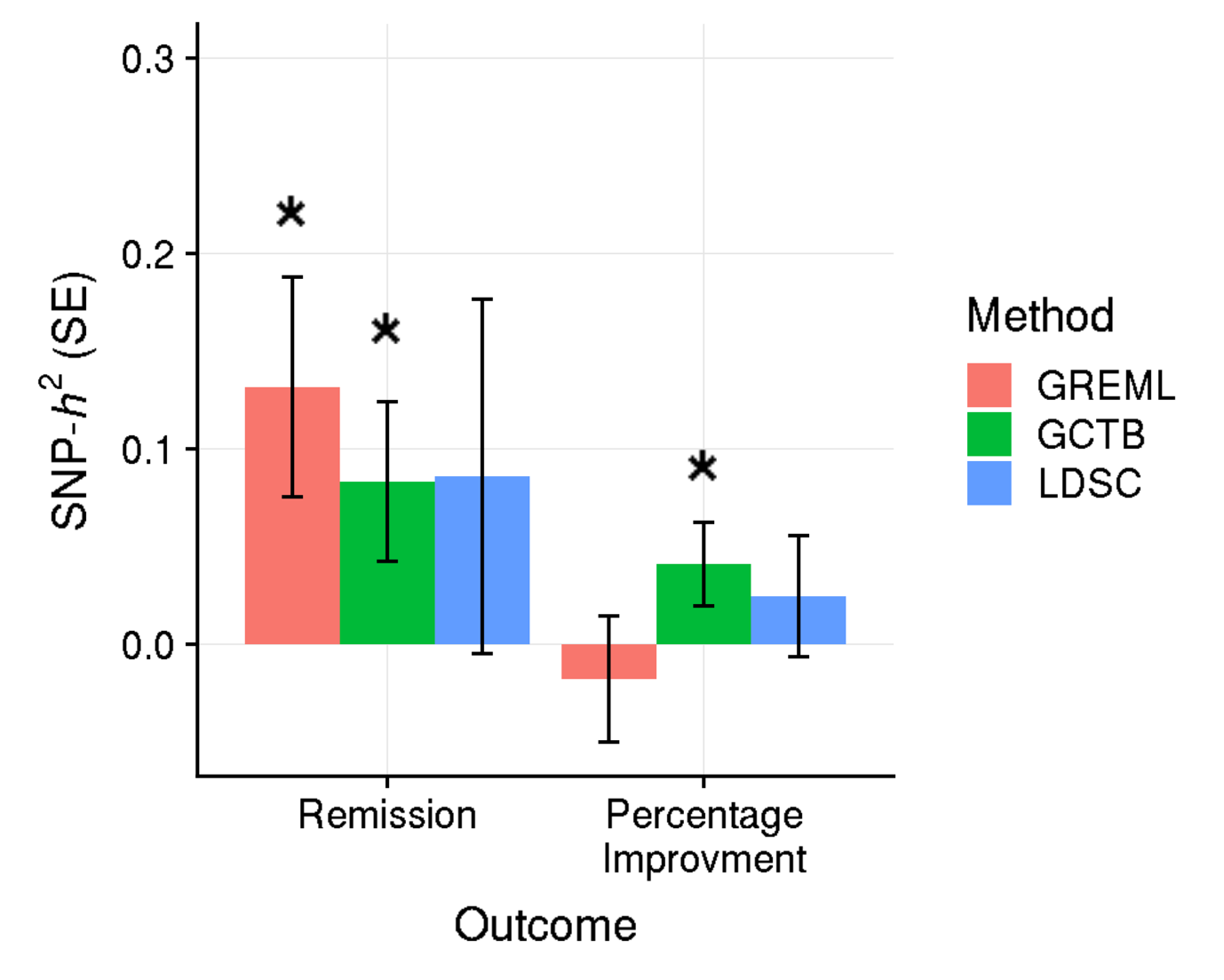

Figure 18. SNP-based heritability estimates for Remission and Percentage Improvement.

*Note. Figure shows across sample estimates of SNP-based heritability, estimated using (mega-) GREML, GCTB and LDSC. * indicates estimate is significantly different from zero, at p<0.05.*

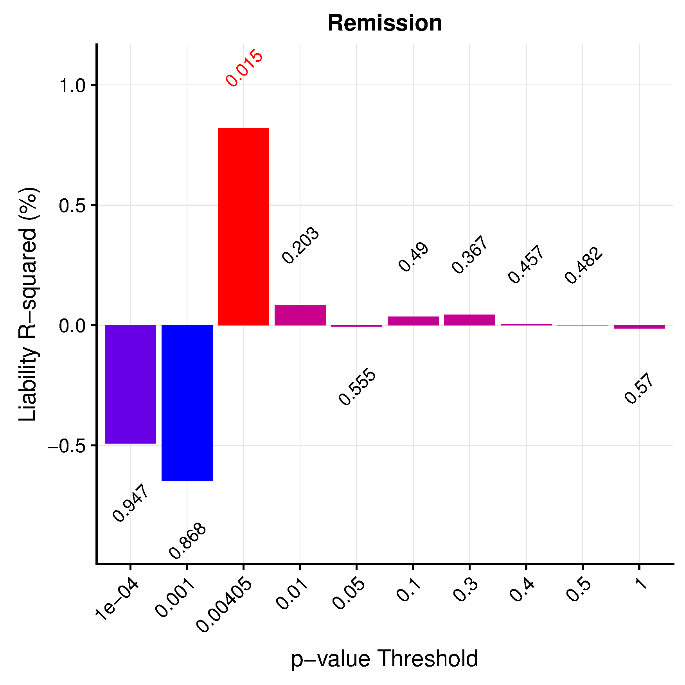

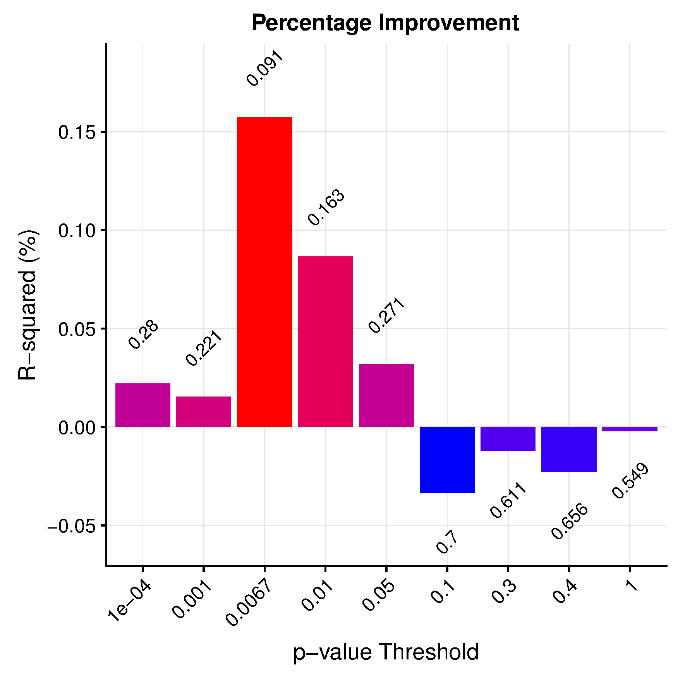

Figure 19. Meta-analysis of Remission and Percentage Improvement polygenic score associations with Remission and Percentage Improvement across prospectively-assessed cohorts.

Note. One-sided p-values are shown above or below each p-value threshold bar, with p-values <0.05 highlighted in red. Figure shows 9 standard p-value thresholds, and the optimal p-value threshold. R-squared estimates are signed to indicate the direction of the association.

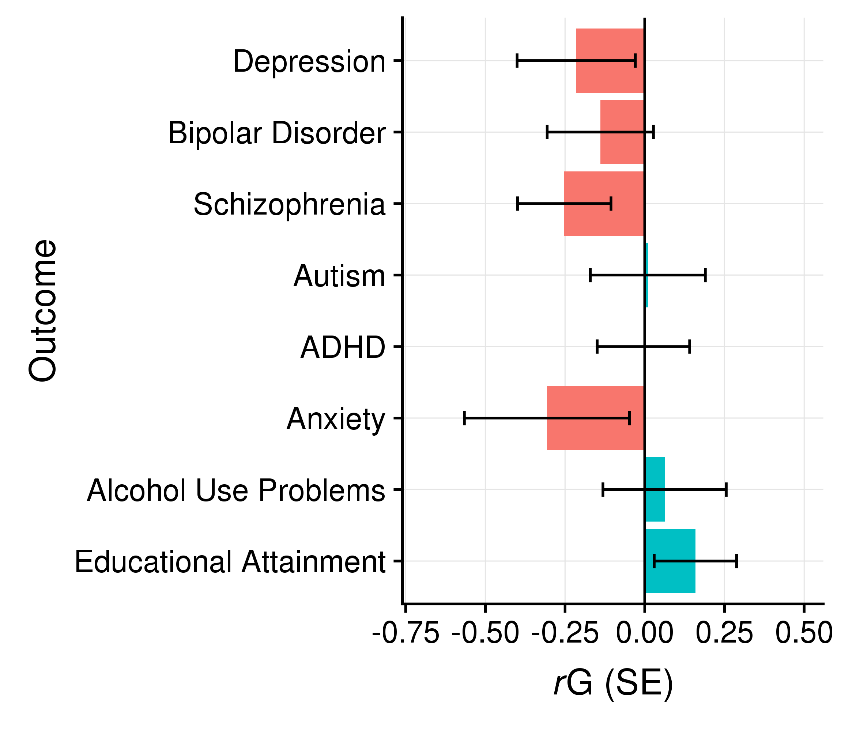

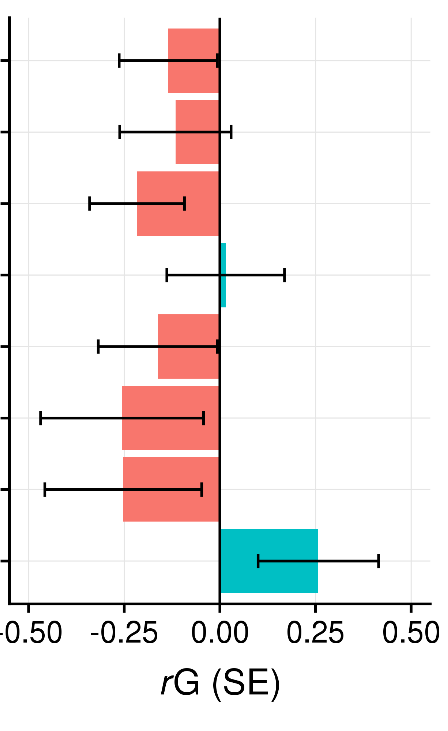

Percentage Improvement

Remission

Figure 20. LDSC genetic correlation estimates between antidepressant response and mental health phenotypes.

*Note. Estimates are based on a constrained intercept of 1. Unconstrained estimates were consistent for Remission but could not be completed for Percentage Improvement due to a negative SNP-based heritability estimate.*

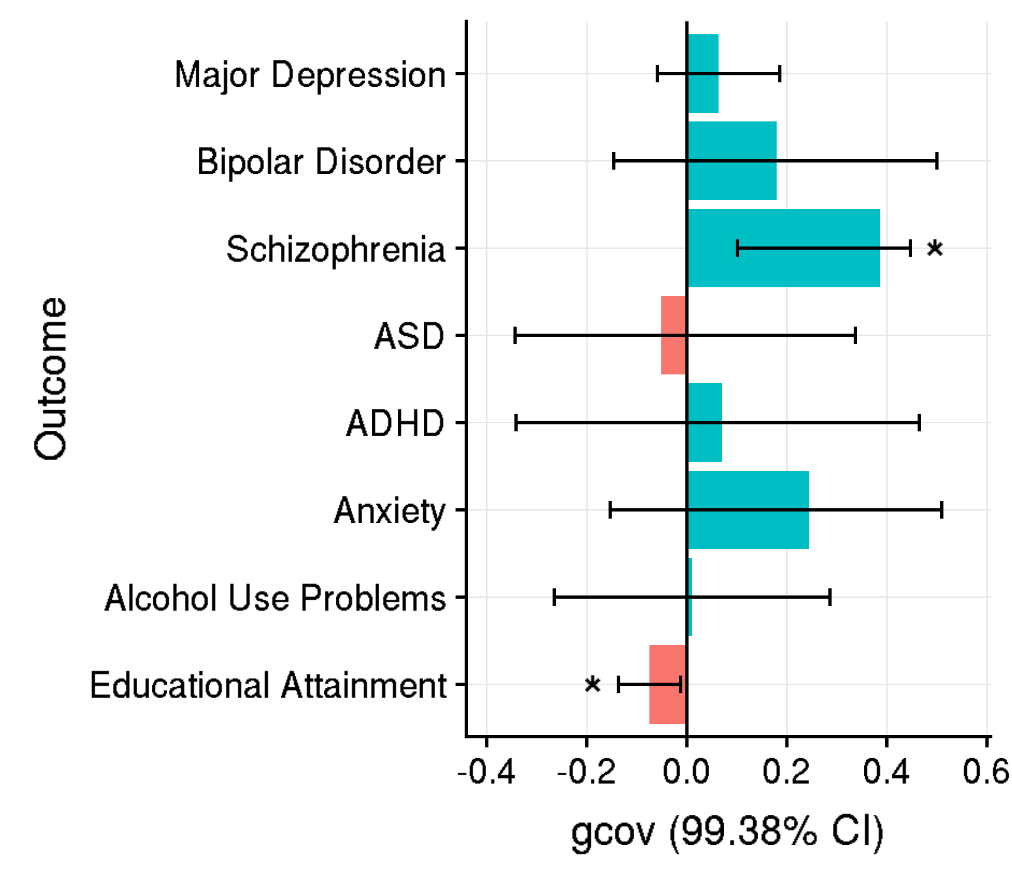

Figure 21. Genetic covariance estimates between TRD in Generation Scotland and mental health phenotypes from AVENGEME analysis

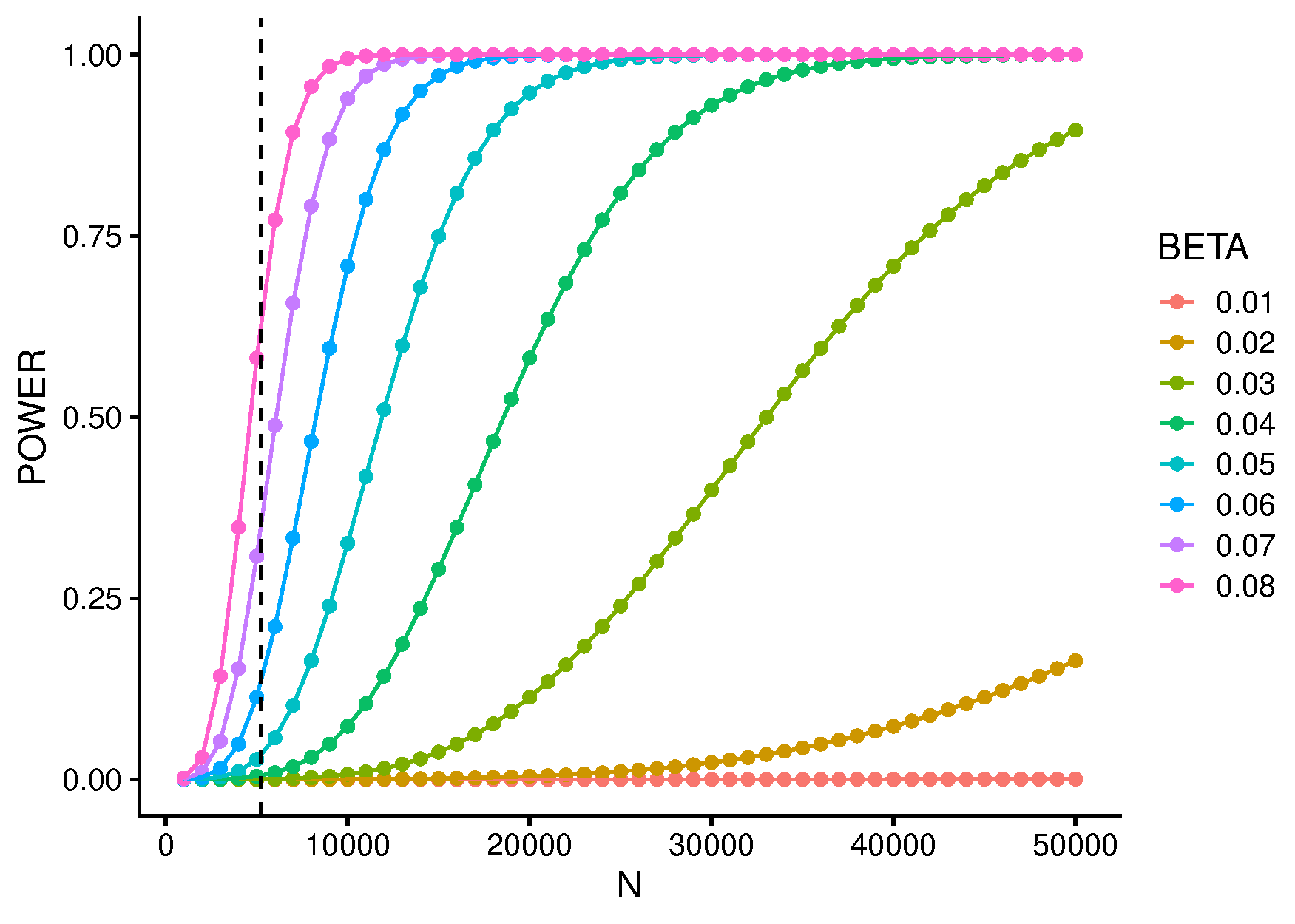

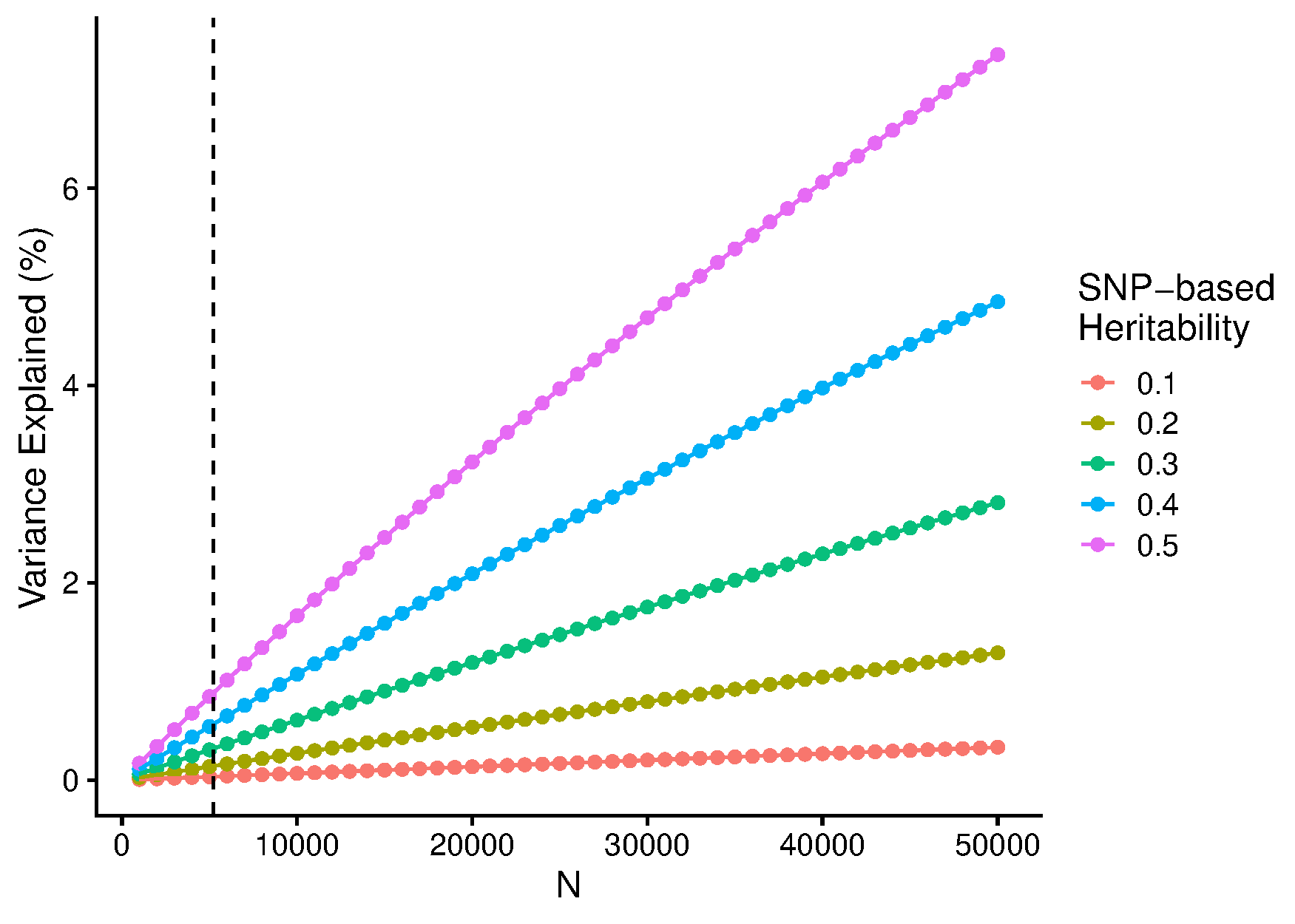

**A)**

**B)**

Figure 22. Power and predictive utility of GWAS.

Note. A) Power to detect genetic variants of varying effect size at genome-wide significance. Power calculation is based on a quantitative outcome. The dash vertical line indicates the sample size of the current Percentage Improvement GWAS. B) Variance explained (*R*^2^) by polygenic scores derived using quantitative traits with varying SNP-based heritability. The dash vertical line indicates the sample size of the current Percentage Improvement GWAS.

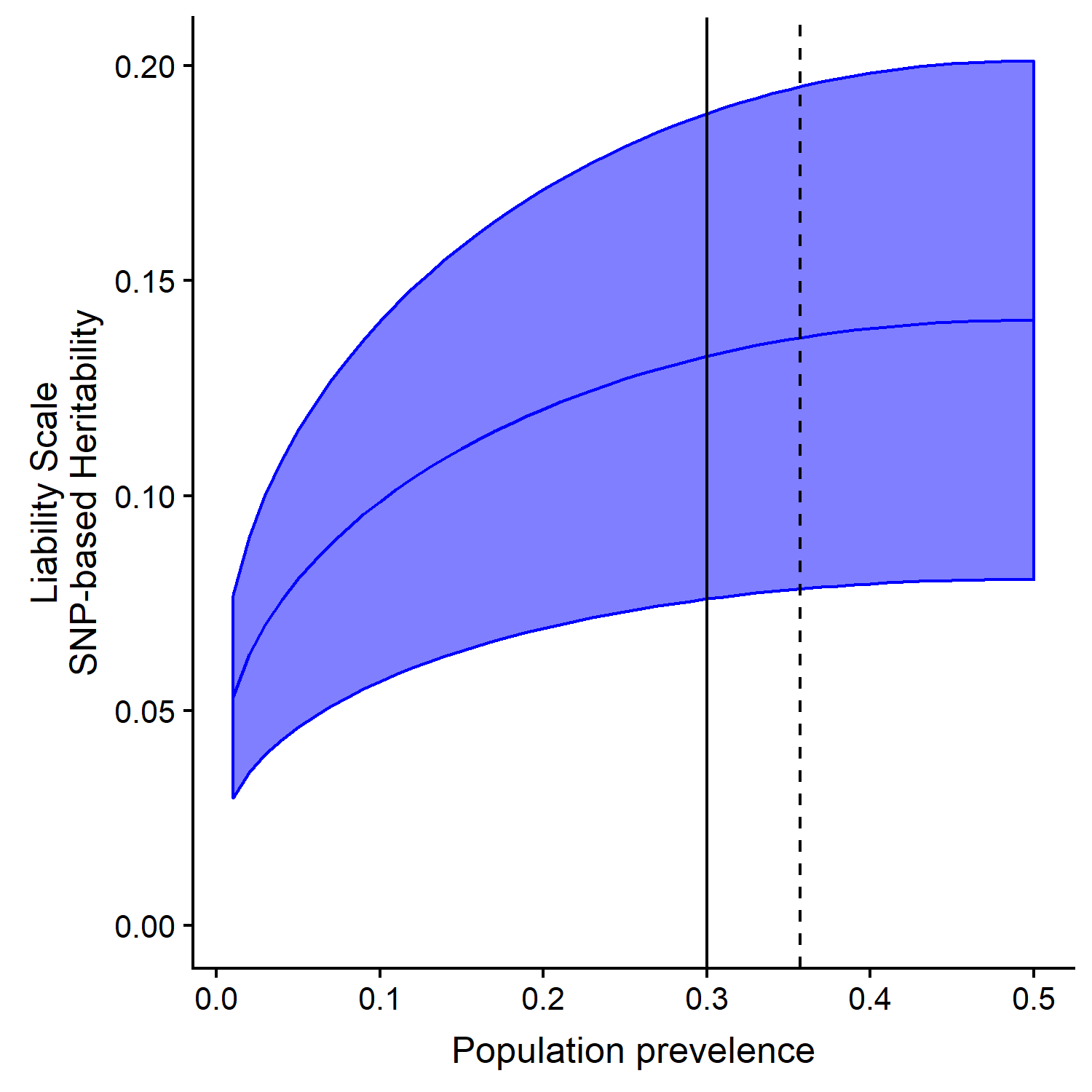

Figure 23. Liability scale SNP-based heritability of Remission across a range of population prevalence.

Note. The observed SNP-based heritability estimate used in this figure was calculated using Mega-GREML. The solid line indicates a population prevalence of 0.3, and the dashed line indicates the observed proportion of remitters in the sample (0.357). The blue shaded area indicates the standard error of the liability scale SNP-based heritability.

References**:**

11.	Wing JK, Sartorius N, Üstün TB. *Diagnosis and Clinical Measurement in Psychiatry: A Reference Manual for SCAN*. Cambridge University Press; 1998.
